# Brain, genetic and demographic factors predict current body fat estimate and weight gain in (pre)adolescents: evidence from the ABCD study

**DOI:** 10.64898/2026.06.25.26356585

**Authors:** Ilkka Suuronen, Jetro J. Tuulari, Ru Li, Ashmeet Jolly, Harri Merisaari, Antti Airola, Hilyatushalihah K. Audah, Aaron Barron, Niloofar Hashempour, Silja Luotonen, Elmo P. Pulli, Aylin Rosberg, Minna Kyläniemi, Riina Kaukonen, Riikka Lund, Esko Pakarinen, Hasse Karlsson, Riikka Korja, Jakob Seidlitz, Richard A.I. Bethlehem, Isabella L.C. Mariani-Wigley

## Abstract

**IMPORTANCE:** Childhood obesity is a growing global health concern associated with adverse physical, psychiatric, and neurodevelopmental outcomes. Although previous neuroimaging studies have linked obesity to widespread alterations in brain structure and function, it remains unclear how well multimodal neuroimaging measures and genetic markers can predict future weight gain and inform early intervention strategies.

**OBJECTIVE:** To evaluate the predictive utility of multimodal MRI measures and polygenic risk scores for obesity in estimating proportional body weight at baseline and predicting weight gain over one year in preadolescent children.

**DESIGN, SETTING, AND PARTICIPANTS:** This study used data from the Adolescent Brain Cognitive Development (ABCD) Study, a large-scale, multisite longitudinal cohort of children aged 9 to 10 years (N = 11,880).

Analyses included baseline data collected between 2016 and 2018, and one-year follow-up data collected between 2018 and 2020 across multiple imaging sites.

**MAIN OUTCOMES AND MEASURES:** Elastic net regression models were applied to structural MRI (including diffusion tensor imaging) and resting-state functional MRI data to predict baseline triponderal mass index (TMI), a weight-for-height measure that more accurately reflects adiposity in children than body-mass index (BMI). Longitudinal classification models were developed to predict excess weight gain relative to normative developmental trajectories at one-year follow-up. Models were evaluated with and without the inclusion of polygenic risk scores and other non-imaging covariates. Generalizability was assessed using leave-one-site-out cross-validation.

**RESULTS:** Structural MRI measures predicted baseline TMI with an R² of 0.21, whereas resting-state functional MRI measures predicted TMI with an R² of 0.08. Classification models predicted one-year weight gain with area under the receiver operating characteristic curve (AUC) values of 0.73 for structural MRI and 0.60 for resting-state functional MRI. Including polygenic risk scores and other covariates improved model performance (structural MRI: R² = 0.25, AUC = 0.75; resting-state functional MRI: R² = 0.15, AUC = 0.69). Leave-one-site-out cross-validation revealed reduced generalizability across imaging sites (structural MRI R² = 0.13–0.17; resting-state functional MRI R² = 0.02–0.09; structural MRI AUC = 0.73–0.74; resting-state functional MRI AUC = 0.60–0.67).

**CONCLUSIONS AND RELEVANCE:** Multimodal MRI measures were associated with proportional body weight and demonstrated modest predictive utility for future weight gain in preadolescent children, explaining up to one fifth of the variance in weight-related outcomes. The addition of genetic and non-imaging variables improved prediction accuracy, underscoring the multifactorial nature of childhood obesity. However, the observed decline in performance under site-wise cross-validation highlights the need to address site-related variability to enhance reproducibility and generalizability in neuroimaging-based predictive models of pediatric obesity.

## 1 Introduction

Pediatric obesity and overweight are pressing global public health challenges that demand urgent action due to their alarming rise and significant long-term consequences.^1^ In 2022, more than 390 million children and adolescents aged 5–19 years were overweight, including 160 million who were living with obesity, with the prevalence of obesity in developmental ages having nearly tripled since 1990.^2^ Children who are overweight or obese are more likely to remain so into adulthood, thereby increasing their risk of cardiometabolic disease, certain cancers, and premature mortality^3^, as well as psychiatric disorders such as depression, anxiety, and eating disorders.^4^ Collectively, these adverse outcomes reduce quality of life for affected individuals and families and place a substantial burden on healthcare systems, underscoring the need for effective preventive interventions and improved treatment strategies.^1^ Despite this urgency, research on pediatric obesity has been hampered by inconsistent findings—particularly in neuroimaging studies—alongside methodological limitations including small sample sizes, the use of imprecise weight measures, and limited integration of genetic factors. This study aims to address these gaps through a comprehensive, multidimensional investigation of pediatric obesity.

The etiology of childhood obesity is complex and multifactorial, arising from interactions between biological factors such as genetic predisposition, physiological regulation, and brain structure and function, as well as environmental influences including diet, physical activity, and social context. The development of obesity involves not only a sustained positive energy balance but also an upward shift in the level of adiposity defended by the homeostatic energy balance system, which helps explain why weight lost through dietary or lifestyle interventions is often regained. Thus, obesity is not best understood as a passive accumulation of excess calories, but rather as a disorder of energy homeostasis.^5^ In addition to homeostatic mechanisms, non-homeostatic processes such as reward-driven hedonic eating can promote excess energy intake and drive body weight above the level defended by homeostatic regulation. Under typical conditions, the homeostatic system regulates the body weight effectively, compensating for such non-homeostatic influences; however, persistent reward-driven overeating can overwhelm this regulatory capacity, contributing to the development and maintenance of obesity.^6^ These complementary pathways to excess weight gain are comprehensively reviewed by Duis and Butler (2022).^7^

Structural and functional magnetic resonance imaging (MRI) studies have linked childhood obesity to differences in brain anatomy and function. Structural MRI studies have reported associations with variation in cortical thickness,^8^ total gray matter volume,^9–14^ and white matter microstructure.^15^ Functional MRI studies have likewise identified alterations in resting-state network connectivity^16,17^ and differences in neural responses to rewarding stimuli, particularly within regions implicated in reward processing^18–22^ and inhibitory control.^20,21,23,24^ Despite these findings, results across studies have been inconsistent.

Reported structural differences have ranged from broad cortical regions, such as the frontal cortex,^8,11,15,25^ to specific subcortical structures including the hippocampus,^26^ pallidum,^27^ amygdala, and nucleus accumbens,^13,14^ yet many of these findings have failed to replicate.^11,27,28^ Similarly, functional MRI studies have yielded mixed results, with some reporting weight-related differences in blood-oxygen-level-dependent (BOLD) responses in the prefrontal and orbitofrontal cortices,^20,21^ inferior frontal gyrus,^24^ insula,^19,29^ and amygdala,^29^ while others have observed no such differences.^30,31^ Moreover, the direction of effects has varied across studies. These inconsistencies complicate the interpretation of neuroimaging findings and limits their translational relevance, underscoring the need for larger, more diverse samples and more standardized analytic approaches in pediatric obesity research.

Individuals with severe obesity frequently experience stigmatization, driven in part by the perception that obesity primarily results from unhealthy lifestyle choices.^32^ However, the underlying heritability of obesity should not be overlooked, as a substantial proportion of individual susceptibility arises from the cumulative effects of numerous common genetic variants.^33^ This susceptibility is largely explained by a polygenic model, characterized by the combined effects of many variants with small individual impacts. To apply this model effectively, a score that reflects the combined impact of these multiple variants on inherited susceptibility is needed. The genome-wide polygenic score (GPS) meets this need by consolidating all prevalent variants into a unified quantitative indicator of inherited susceptibility.^34^ Recently, a polygenic predictor comprising 2.1 million common variants to quantify this susceptibility was derived and validated on 300,000 individuals with ages spanning from birth to middle age.^35^

During adolescence, body weight is not proportional to height squared, the assumption underlying body mass index (BMI). For this reason, BMI z-scores are typically used for children and adolescents. The triponderal mass index (TMI) is an indirect measure of body adiposity, computed as body mass in kilograms divided by height in meters cubed.^36^ Compared with BMI z-scores, TMI has been shown to have a lower rate of obesity misclassification, to provide better estimates of body fat levels, and to remain approximately constant throughout adolescence.^36^ Subsequent studies have confirmed that TMI is superior to BMI in predicting body fat percentage,^37–39^ although it is less strongly correlated with fat mass.^37^ For the classification of obesity or high adiposity, TMI is considered more effective^37–39^ or at least as effective as BMI.^40^ Because of its relative stability during adolescence and its improved performance in identifying high adiposity, we considered TMI preferable to BMI for the purposes of this study.

In this study, we partially replicate and substantially extend earlier work by Adise et al.^41^ by conducting a multimodal, cross-sectional and longitudinal, machine-learning–based investigation of associations between brain measures and body mass in children. Using the updated ABCD Study data release 5.0, we (1) performed cross-sectional prediction of baseline body mass from structural MRI and resting-state fMRI data; (2) conducted longitudinal prediction of weight gain versus stable weight status at one-year follow-up; (3) assessed model generalizability using leave-one-site-out cross-validation; (4) incorporated polygenic risk scores for obesity to account for genetic susceptibility; and (5) applied auxiliary principal component analysis (PCA) to reduce the dimensionality of neuroimaging features and identify components capturing major patterns in the data.

## 2 Methods

### 2.1 Study design

Data for this study were obtained from the openly available Adolescent Brain Cognitive Development (ABCD) Study® data release 5.0.^42^ The ABCD Study is a multisite, 10-year longitudinal cohort study that began in late childhood, with 11,880 participants enrolled at ages 9–10 years (baseline assessment), with the aim of investigating brain and cognitive development through adolescence. Assessments are performed in the laboratory each year, while MRI data are acquired every two years. The ABCD dataset includes multimodal neuroimaging data, including structural, diffusion, task-based, and resting-state functional MRI, as well as measures of social, emotional, and cognitive development, physical and mental health, and environmental factors. This study used the tabular data release, which contains derived measures for each neuroimaging modality.

### 2.2 Sample exclusion criteria

The exclusion criteria for participation in the ABCD study have been reported previously.^43^ Consistent with our aim to replicate and extend the work of Adise et al.^41^, we applied the same expert-informed sample exclusion criteria to both baseline and one-year follow-up data as specified in their study.

#### 2.2.1 Baseline sample

For the baseline sample, participants were excluded if they met any of the following criteria: (1) missing or clearly erroneous anthropometric values (e.g., height = 6.5 inches); (2) very low body weight (sex-specific TMI < 5th percentile); (3) use of medications known to affect food intake; or (4) presence of psychiatric disorders, neurological or intellectual disabilities, or eating disorders (current or in partial remission). Following the methodology of the original study being replicated, participants were additionally excluded if they reported being transgender, had mislabeled sex information, or provided inconsistent sex-specific pubertal development data. Only participants with complete demographic information—including age, sex, pubertal status, race, and maternal education—were retained.

After applying these criteria, the final baseline sample comprised participants with valid structural T1-weighted MRI (N = 8,999), diffusion tensor imaging (DTI; N = 8,234), and resting-state fMRI (N = 7,639) data (**Figure 1**).

**Figure 1.**
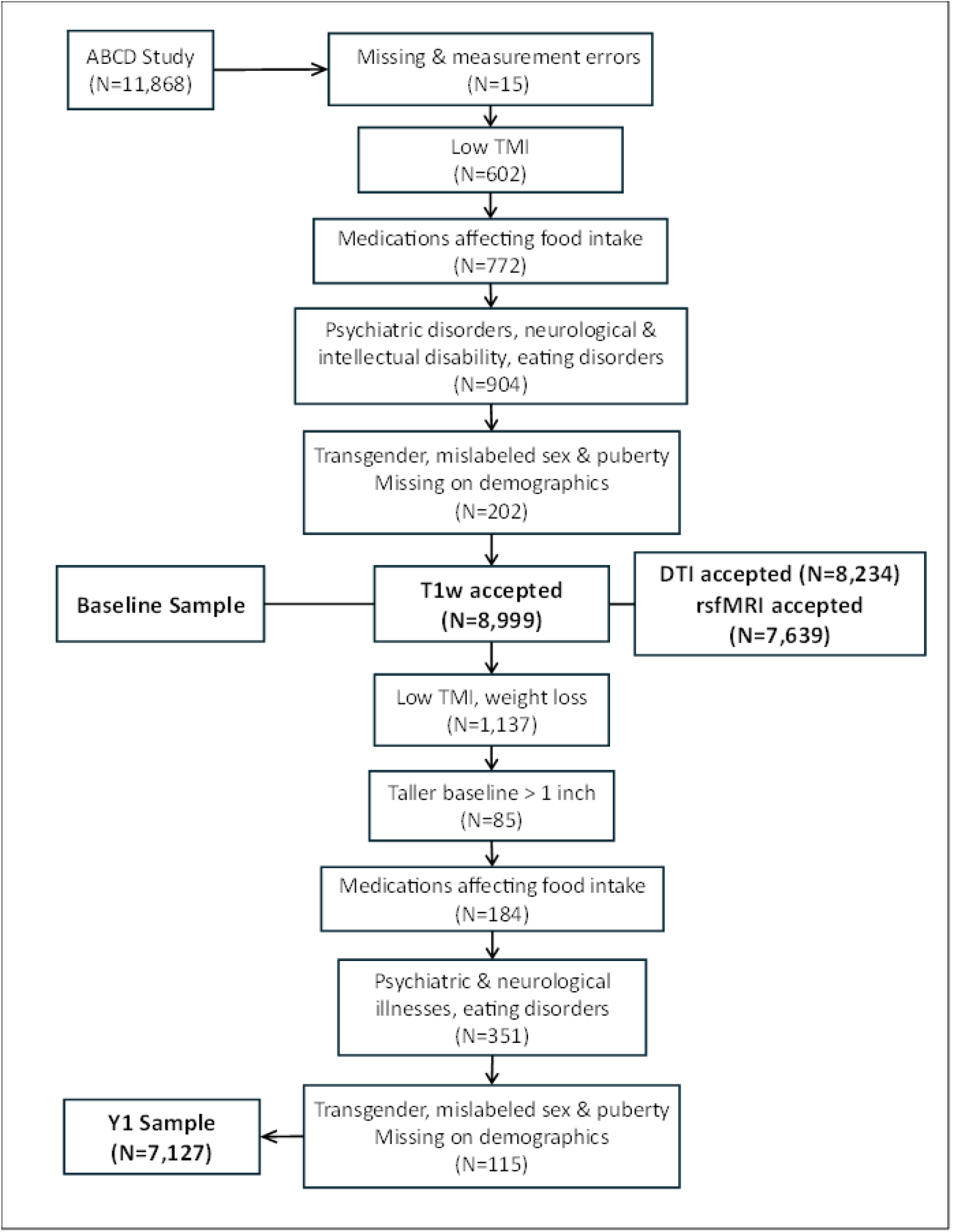
A flow chart of participant exclusions. TMI = Triponderal mass index, DTI = Diffusion tensor imaging, rsfMRI = resting-state functional magnetic resonance imaging. T1w = T1-weighted structural magnetic resonance imaging.

#### 2.2.2 Follow up data one year after the baseline (Y1 sample)

Starting from the baseline sample with accepted T1-weighted imaging data (N = 8,999), additional exclusions were applied for the one-year follow-up analysis based on the following criteria: (1) very low body weight (sex-specific TMI < 5th percentile); (2) weight loss relative to baseline; (3) a decrease in height exceeding one inch compared with baseline; (4) use of medications affecting food intake; and (5) presence of psychiatric disorders, neurological or intellectual disabilities, or eating disorders (current or in partial remission). Participants who reported being transgender, had mislabeled sex information, inconsistent sex-specific pubertal data, or incomplete demographic information were also excluded.

Following these exclusions, the final one-year follow-up (Y1) sample comprised 7,127 children (**Figure 1**).

#### 2.2.3 Adiposity changes between baseline and Y1

In line with the approach used by Adise et al.^41^ changes in adiposity between baseline and the one-year follow-up (Y1) were categorized into two groups. The weight gain (WG) group was defined as participants with a TMI change z-score ≥ 0.2, together with either a weight increase greater than 1 SD or classification as overweight (TMI ≥ 85th percentile) at Y1.

The weight stable (WS) group comprised participants with a TMI change z-score between −0.2 and 0.2 who remained within the normal-weight range (TMI < 85th percentile) at both time points. The WG group included 1,017 participants, while the WS group included 2,085 participants. Figure 2 shows TMI at baseline plotted against TMI at Y1 for all participants (Figure 2A), as well as separately for the WG and WS groups (Figure 2B).

**Figure 2.**
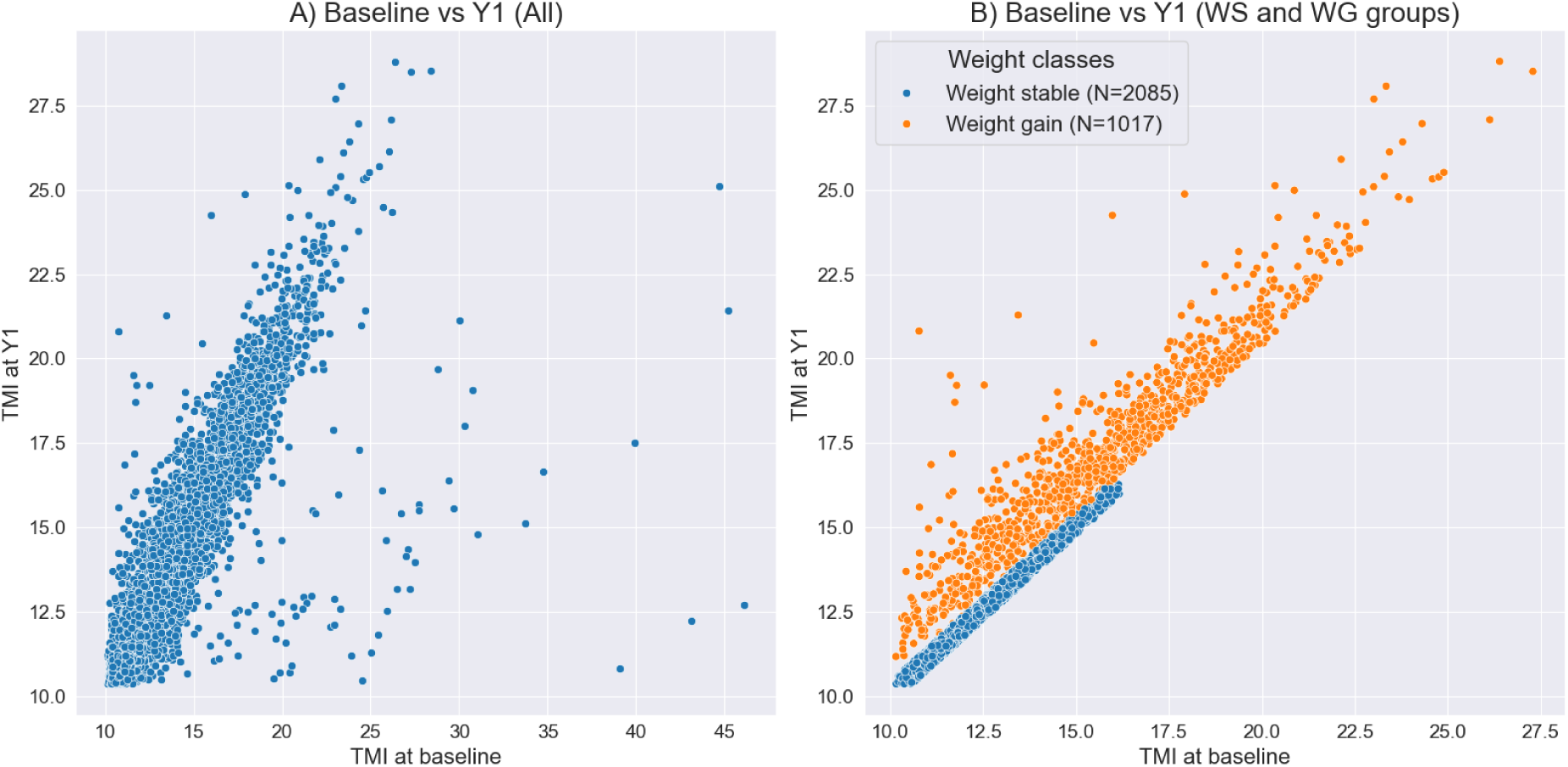
TMI distributions. A) TMI at one year follow-up plotted against baseline for all participants. B) TMI at one year follow-up plotted against baseline for weight gain and weight stable groups.

Given that children naturally gain weight as they develop, this study specifically focused on prominent weight gain over a relatively short period, which can have important health implications and may signal early preclinical eating disorders. Within this sample, a notable proportion of children exhibited weight gain exceeding normative developmental trajectories. The study therefore aimed to investigate potential brain correlates that distinguish children with excessive weight gain from those with developmentally typical changes. Characterizing these neural differences may provide insight into the neurobiological mechanisms underlying preclinical eating disorders and inform early interventions targeting pathological overeating.

### 2.3 Imaging measures

Multimodal brain imaging data were acquired on 3T scanners from three major manufacturers (Siemens, General Electric, and Philips) across 21 imaging sites. Structural T1-weighted (T1w) images were acquired using a three-dimensional inversion-prepared gradient-echo sequence, with prospective motion correction when available, and a voxel size of 1 mm isotropic. Diffusion MRI (dMRI) data were acquired using a multiband echo-planar imaging (EPI) sequence with a slice acceleration factor of 3, comprising 96 diffusion-weighted directions, seven b = 0 volumes, and four b-value shells (6 directions at b = 500 s/mm², 15 directions at b = 1,000 s/mm², 15 directions at b = 2,000 s/mm², and 60 directions at b = 3,000 s/mm²), with a voxel size of 1.7 mm isotropic. Functional MRI (fMRI) data were acquired using multiband EPI with a slice acceleration factor of 6, a voxel size of 2.4 mm isotropic, and a repetition time (TR) of 800 ms. For both dMRI and fMRI acquisitions, field map scans were collected to enable correction of B0-related geometric distortions.

#### 2.3.1 Structural brain imaging measures

Structural brain imaging data were parcellated using the Destrieux atlas^44^ for cortical regions and the FreeSurfer subcortical atlas^45^ for subcortical regions of interest. Structural measures included cortical thickness, cortical surface area, and volumes of subcortical structures.

Diffusion tensor imaging (DTI) measures of fractional anisotropy (FA) and mean diffusivity (MD) were estimated for white matter adjacent to cortical regions defined by the Destrieux atlas, as well as for subcortical structures.

#### 2.3.2 Resting-state functional brain imaging measures

Resting-state functional brain imaging data were parcellated using the Gordon atlas^46^ and grouped into canonical Gordon networks. Average pairwise correlations between regions within each network, as well as between cortical networks and subcortical regions, were computed and used as measures of functional connectivity in the analyses.^47^

### 2.4 Non-imaging measures

Non-imaging measures included as covariates in the analyses were participants’ age at the time of interview, scanner model, sex, ethnicity, handedness, pubertal status, and highest parental education, as well as polygenic risk scores for obesity. Puberty estimates were derived as the mean of child- and parent-reported measures. Participants’ triponderal mass index (TMI), calculated as body mass in kilograms divided by height in meters cubed, was used as the target variable in the machine-learning–based regression analyses.

### 2.5 Analytic approach

#### 2.5.1 Machine learning

The machine-learning approach was implemented following the methodology described by Adise et al.^41^ Two sets of analyses were conducted: (i) cross-sectional prediction of baseline TMI (ages 9–10 years) using derived brain measures and non-imaging covariates as predictors, and (ii) longitudinal prediction of weight gain versus stable weight status at one-year follow-up using baseline derived brain measures and sociodemographic covariates as predictors. Both regression and classification analyses were performed separately for structural and functional brain measures, and models were evaluated both with and without the inclusion of non-imaging covariates.

An elastic net model was used for all machine-learning analyses. For the classification task, a logistic link function was applied to model outcome probabilities. Nested cross-validation was used to optimize model hyperparameters, including the overall regularization strength and the ratio of L1 and L2 penalty terms. Prior to model fitting, additional participant exclusions were applied for missing target variable values, non-singleton participants, and failed quality control. The prediction pipeline consisted of the following steps: (1) imputation of missing values (median imputation for numeric variables and most-frequent value imputation for categorical variables); (2) dummy encoding of categorical variables; (3) z-score standardization; and (4) model fitting and prediction.

To replicate the analyses reported by Adise et al.^41^ for predicting baseline TMI and weight gain versus stable weight status from brain imaging measures and non-imaging covariates, we used 5-fold cross-validation. To assess generalizability across imaging sites, we additionally employed leave-one-site-out cross-validation^48–50^, in which data from each site were held out in turn as an independent test set while models were trained on data from all remaining sites. In both validation schemes, nested 10-fold cross-validation within the training data was used to optimize model hyperparameters. Model performance was evaluated using the coefficient of determination (R²) for regression analyses and the area under the receiver operating characteristic curve (AUC) for classification analyses.

Performance metrics were averaged across test folds and, for leave-one-site-out analyses, additionally reported separately for each held-out site.

The analyses were performed on Python version 3.12.11^51^ with external libraries NumPy 1.26.4^52^, Pandas 2.3.2^53^ and Scikit-learn 1.7.2^54^.

#### 2.5.2 Conventional statistics with dimension reduction

Principal component analysis (PCA) was employed to reduce the dimensionality of the brain imaging data and to identify components representing key patterns within the dataset. Given the large number of variables included in the analyses, the conventional criterion of retaining components with eigenvalues greater than 1 (Kaiser’s rule) was deemed impractical. Instead, components were selected based on the inflection point of the scree plot, corresponding to the point at which additional components contributed diminishing gains in explained variance.

When scree plots were difficult to interpret, component selection further considered both the variance explained by individual components and the cumulative explained variance.

Although no universally accepted thresholds exist, priority was given to components explaining more than 5% of the variance individually and/or contributing to a cumulative explained variance exceeding 30%. Varimax rotation was applied to improve the interpretability of the principal components (PCs). This approach was chosen to accommodate the inherent complexity of neuroimaging data, in which total explained variance is typically lower than the conventional 50% benchmark often used in less complex datasets.

To assess adiposity-related neural correlates, two complementary meta-analytic frameworks were implemented. First, cross-sectional associations were examined using linear regression models, with baseline TMI as the dependent variable and PCA-derived neuroimaging components as independent variables. Second, longitudinal adiposity change was modeled using logistic regression, with weight trajectory as a binary outcome (0 = weight stable, 1 = weight gain). All models adjusted for age, pubertal stage, race, and maternal education.

Site-specific regression estimates were pooled using random-effects meta-analysis models to account for between-site heterogeneity. Results are reported as effect sizes with 95% confidence intervals (CIs) for linear models and as odds ratios (ORs) with 95% CIs for logistic models.

Both frameworks incorporated subgroup analyses. Meta-analyses were conducted with subgroup analysis by sex, as well as stratified by scanner type with sex included as a covariate. Stratification by sex and scanner type was intended to help distinguish biologically driven heterogeneity (sex-related differences) from technical variability attributable to scanner differences, thereby improving the robustness of the meta-analytic estimates.

Site 22 (N = 21) was excluded because it failed to yield valid parameter estimates during regression modeling, likely due to limited sample size compromising model convergence and stability. In addition, variables associated with high standard errors (SEs) were excluded to enhance the robustness and reliability of the estimates. These exclusions improved model stability without materially altering the direction or statistical significance of the primary findings.

All PCA and regression analyses were conducted using SPSS version 29.0, while meta-analyses and visualizations were performed in R version 4.4.1.

## 3 Results

### 3.1 Elastic net regression models

We performed two sets of analyses to predict (a) baseline TMI in a cross-sectional regression framework and (b) weight gain beyond normative development at one-year follow-up in a longitudinal classification framework. Both analyses were conducted separately using structural brain imaging measures and resting-state functional brain imaging measures as features and were evaluated both with and without the inclusion of non-imaging covariates. Accordingly, a total of eight machine-learning models were trained and tested using a 5-fold cross-validation strategy for each such scenario, with hyperparameter tuning implemented using nested 10-fold cross-validation. Sample characteristics for each imaging modality and machine-learning task are summarized in Table 1.

**Table 1.**
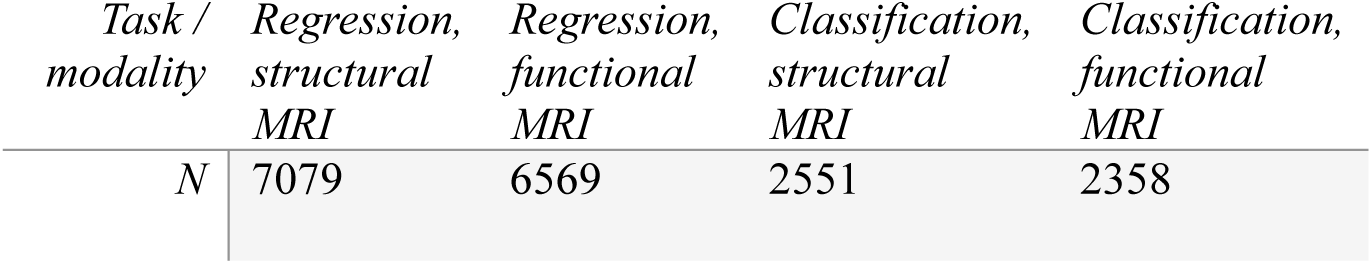
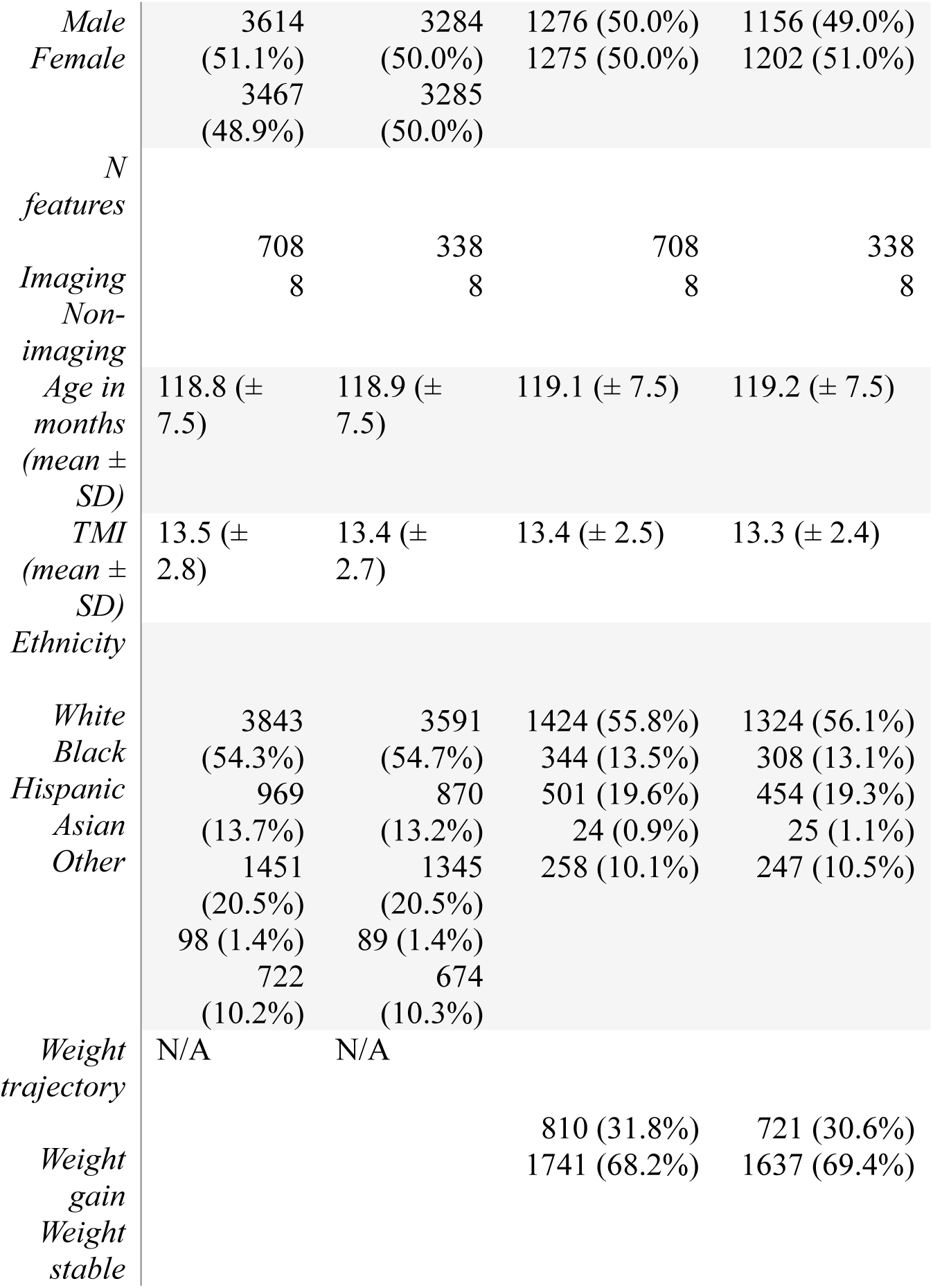
Sample characteristics for each machine learning analysis setting.

#### Structural MRI cross-sectional elastic net regression

When predicting baseline TMI from structural brain measures without non-imaging covariates, the model achieved a mean test R² of 0.21 across cross-validation folds; inclusion of non-imaging covariates increased the mean test R² to 0.25. Training and test performance metrics for cross-sectional baseline TMI prediction are reported in Table 2A. Mean coefficients from the cross-validated elastic net models are projected onto cortical and subcortical surfaces in Figure 3. Structures that cannot be visualized on brain surface plots, as well as non-imaging covariates, are shown in Supplementary Figures S1 and S2. Relative contributions of different predictor categories to the aggregate sum of absolute mean coefficients for models including non-imaging covariates are presented in Supplementary Figure S3.

**Figure 3.**
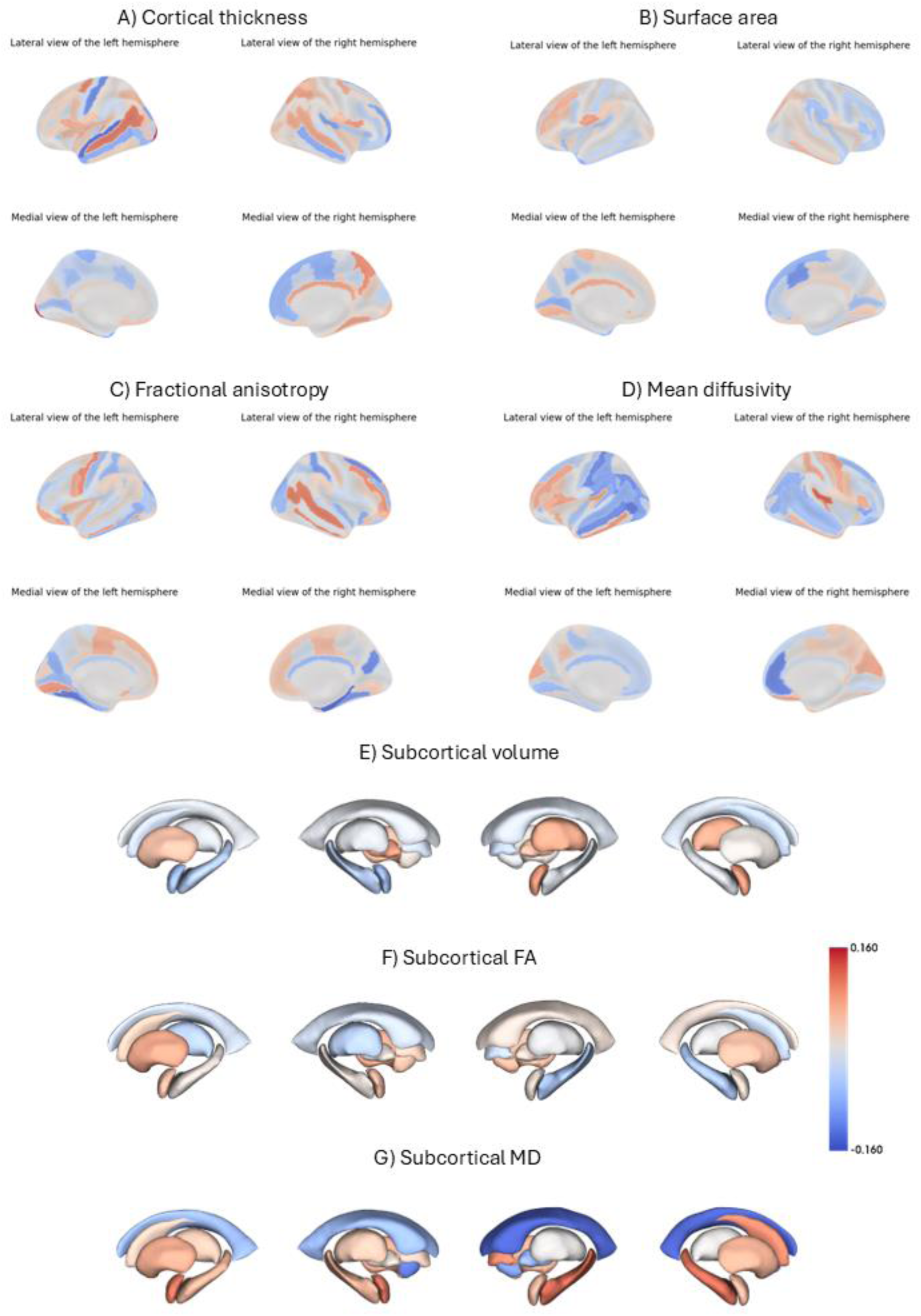
Mean elastic net regression model coefficients across the cross-validation folds for sMRI measures. A) Cortical thickness, B) Cortical surface area, C) White matter fractional anisotropy, D) White matter mean diffusivity, E) Subcortical volume, F) Subcortical fractional anisotropy, G) Subcortical mean diffusivity. Cortical parcellation uses Destrieux atlas and subcortical parcellation uses ASEG subcortical atlas.

**Table 2.**
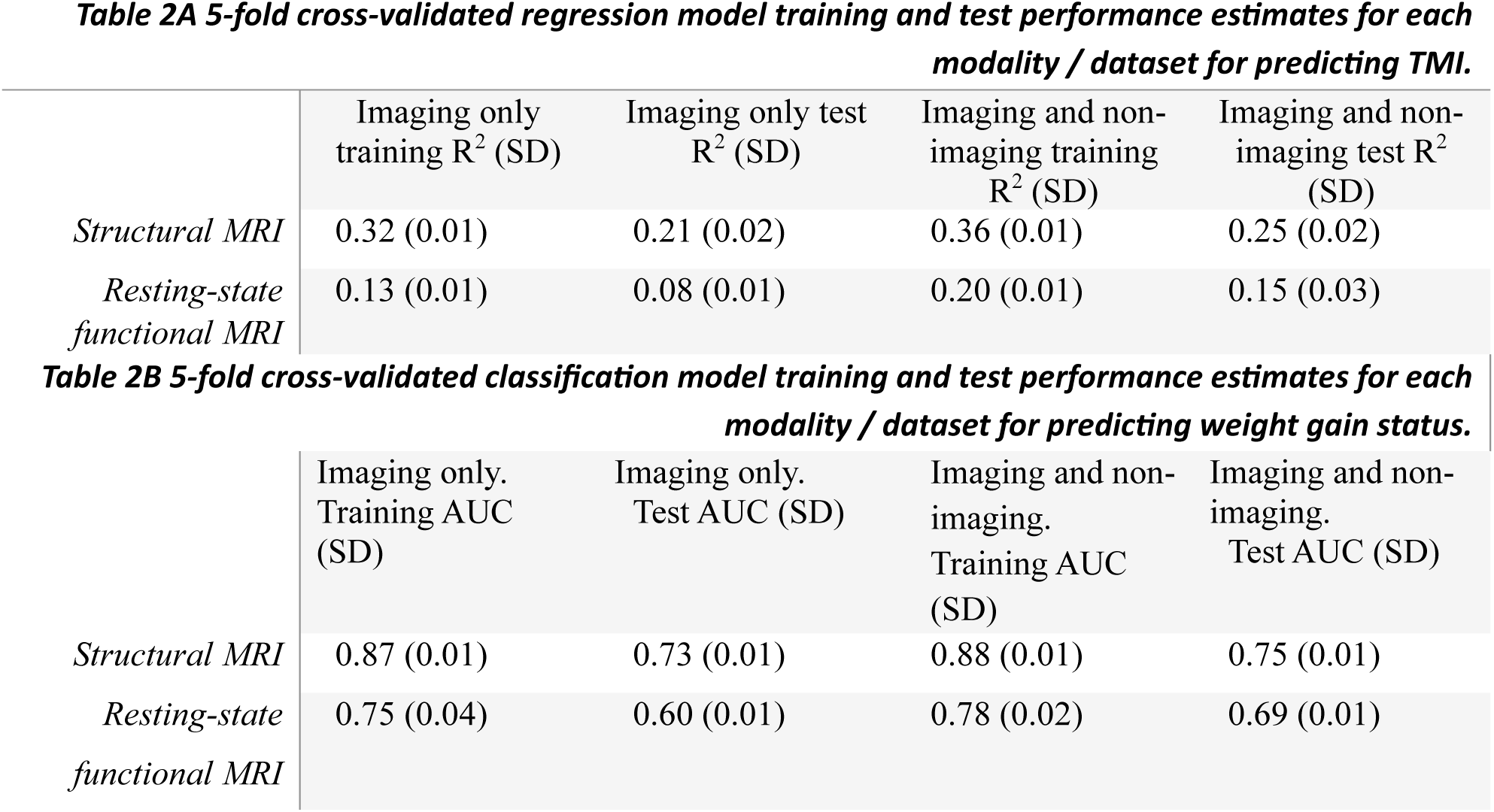
Mean 5-fold cross-validated model performance estimates for regression and classification models across imaging modalities and analytic settings.

**Table 3.** Mean leave-one-site-out cross-validated model performance estimates for regression and classification models using different models and datasets. AUC = Area under curve. SD = Standard deviation.

|  | sMRI imaging features | sMRI and non-imaging features | rs-fMRI imaging features | rs-fMRI and non-imaging features |
| --- | --- | --- | --- | --- |
| *Mean R<sup>2</sup> (SD)* | 0.13 (0.14) | 0.17 (0.13) | 0.02 (0.08) | 0.09 (0.09) |
| *Weighted mean R<sup>2</sup>* | 0.14 | 0.18 | 0.022 | 0.10 |

|  | sMRI imaging features | sMRI and non-imaging features | rs-fMRI imaging features | rs-fMRI and non-imaging features |
| --- | --- | --- | --- | --- |
| *mean AUC (SD)* | 0.73 (0.06) | 0.74 (0.06) | 0.60 (0.07) | 0.67 (0.06) |
| *weighted mean AUC* | 0.72 | 0.74 | 0.60 | 0.68 |

Examination of mean model weights across cross-validation folds indicated that T1-weighted MRI–based predictors, including cortical thickness, cortical surface area, and subcortical volumes, accounted for 37.1% of total model weight. Diffusion tensor imaging–based predictors, including fractional anisotropy (FA) and mean diffusivity (MD) of white matter adjacent to cortical regions of interest and subcortical structures, accounted for 51.9% of total model weight. Non-imaging covariates contributed 11.0% of the model’s total weight.

Predictors related to cortical regions of interest generally exhibited the highest individual contributions (6.3%–10.2%), without a clear hemispheric preference. Despite their relatively small number, non-imaging covariates showed comparatively large contributions, with scanner IDs accounting for 6.5% of total model weight. Notably, the polygenic risk score alone accounted for approximately 1.0% of the total model weight.

At the level of individual regions of interest and structural features, the largest positive coefficients were assigned to cerebellar white-matter volume bilaterally, and cortical thickness of the left occipital pole. In contrast, the most negative coefficients were associated with MD of the left cerebellar cortex, volume of the left cerebellar cortex, and FA of white matter adjacent to the right parahippocampal gyrus. Comprehensive lists of mean regression model coefficients and their standard deviations are provided in Online Supplementary Files 1 (imaging data only) and 2 (imaging data with non-imaging covariates).

#### Resting state fMRI cross-sectional elastic net regression

When predicting baseline TMI from resting-state functional connectivity measures without non-imaging covariates, the model achieved a mean test R² of 0.08; inclusion of non-imaging covariates increased the mean test R² to 0.15 (Table 2A). Mean coefficients from the cross-validated elastic net models are visualized as heatmaps of network correlations in Figure 4. Non-imaging covariates are shown in Supplementary Figure S4. Relative contributions of different predictor categories to the aggregate sum of absolute mean coefficients for models including non-imaging covariates are presented in Supplementary Figure S5.

**Figure 4.**
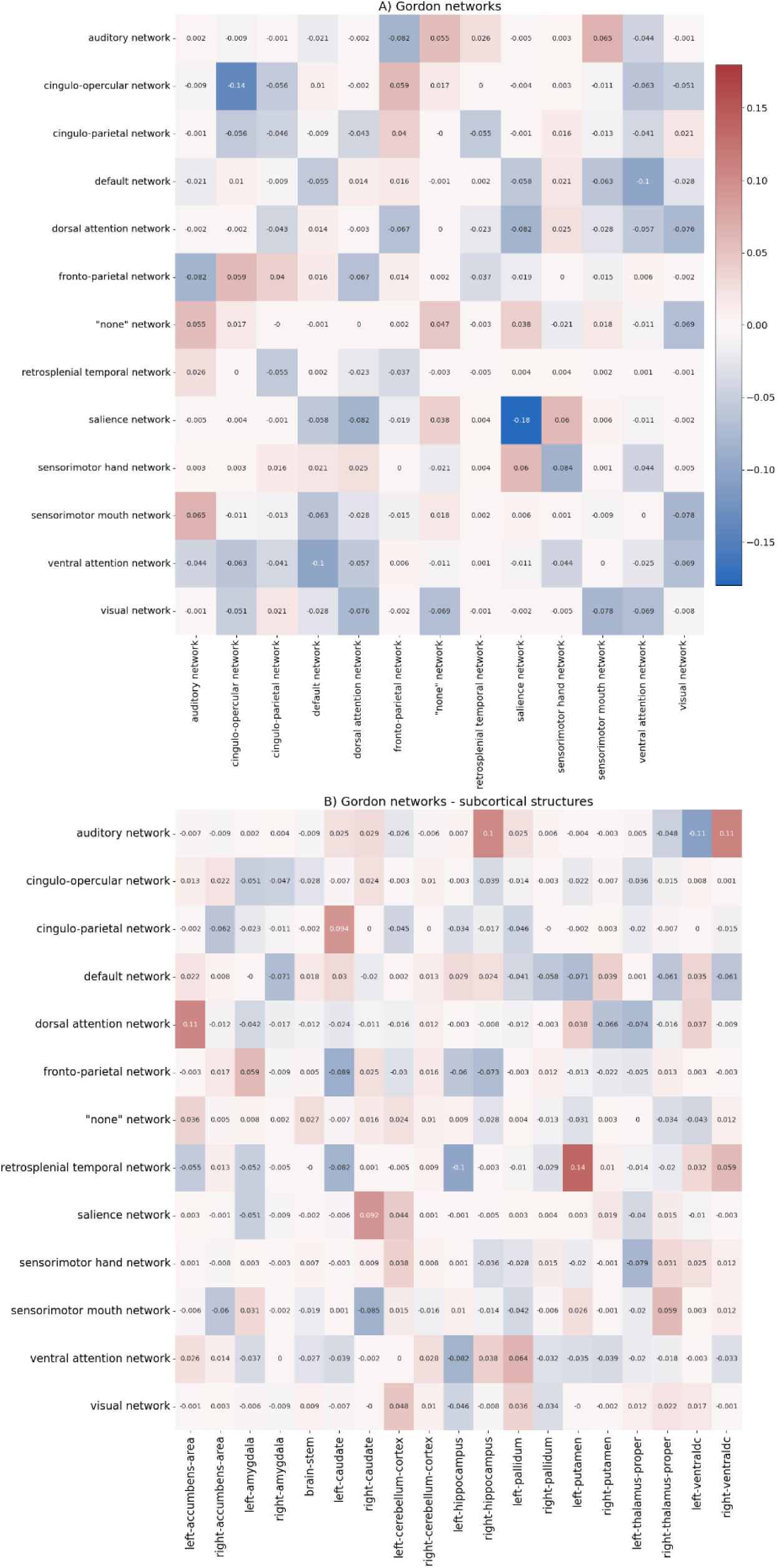
Mean elastic net regression model coefficients for rs-fMRI measures: A) Gordon network intercorrelations, B) Gordon network to subcortex correlations

Connectivity features derived from Gordon network intercorrelations accounted for 22.9% of total model weight, whereas correlations between cortical Gordon networks and subcortical regions accounted for 49.1%, with a left-hemisphere predominance (27.9% left vs. 20.2% right). Non-imaging covariates contributed 28.0% of total model weight, with scanner identifiers accounting for 11.1%, sociodemographic background variables for 13.1%, and polygenic risk scores for 3.7%. The relatively larger contribution of non-imaging covariates in these models may reflect the weaker predictive signal of resting-state network correlation features for baseline TMI, or alternatively the smaller number of rs-fMRI features relative to the total number of structural MRI features, resulting in a greater proportional contribution of non-imaging variables.

At the level of individual connectivity features, the largest positive coefficients were assigned to correlations between the retrosplenial temporal network and left putamen, the dorsal attention network and left nucleus accumbens, the auditory network and right ventral diencephalon, and the auditory network and right hippocampus. Notably, correlations between cortical networks and subcortical regions generally exhibited larger positive coefficients than Gordon network intercorrelations. The largest negative coefficients were associated with within-network correlations in the salience and cingulo-opercular networks, followed by correlations between the auditory network and left ventral diencephalon, the retrosplenial temporal network and left hippocampus, and the ventral attention and default mode networks. Comprehensive lists of mean regression model coefficients and their standard deviations are provided in Online Supplementary Files 3 (imaging data only) and 4 (imaging data with non-imaging covariates).

#### Structural MRI longitudinal classification of future weight gain

When predicting weight trajectory defined as weight gain versus stable weight status at one-year follow-up from baseline structural brain measures without non-imaging covariates, the model achieved a mean test AUC of 0.73; inclusion of non-imaging covariates increased the mean test AUC to 0.75. Training and test performance metrics for longitudinal classification of weight trajectory are reported in Table 2B. Mean coefficients from the cross-validated elastic net models are projected onto cortical and subcortical surfaces in Figure 5. Structures that cannot be visualized on brain surface plots, as well as non-imaging covariates, are shown in Supplementary Figures S6 and S7. Relative contributions of different predictor categories to the aggregate sum of absolute mean coefficients for models including non-imaging covariates are presented in Supplementary Figure S8.

**Figure 5.**
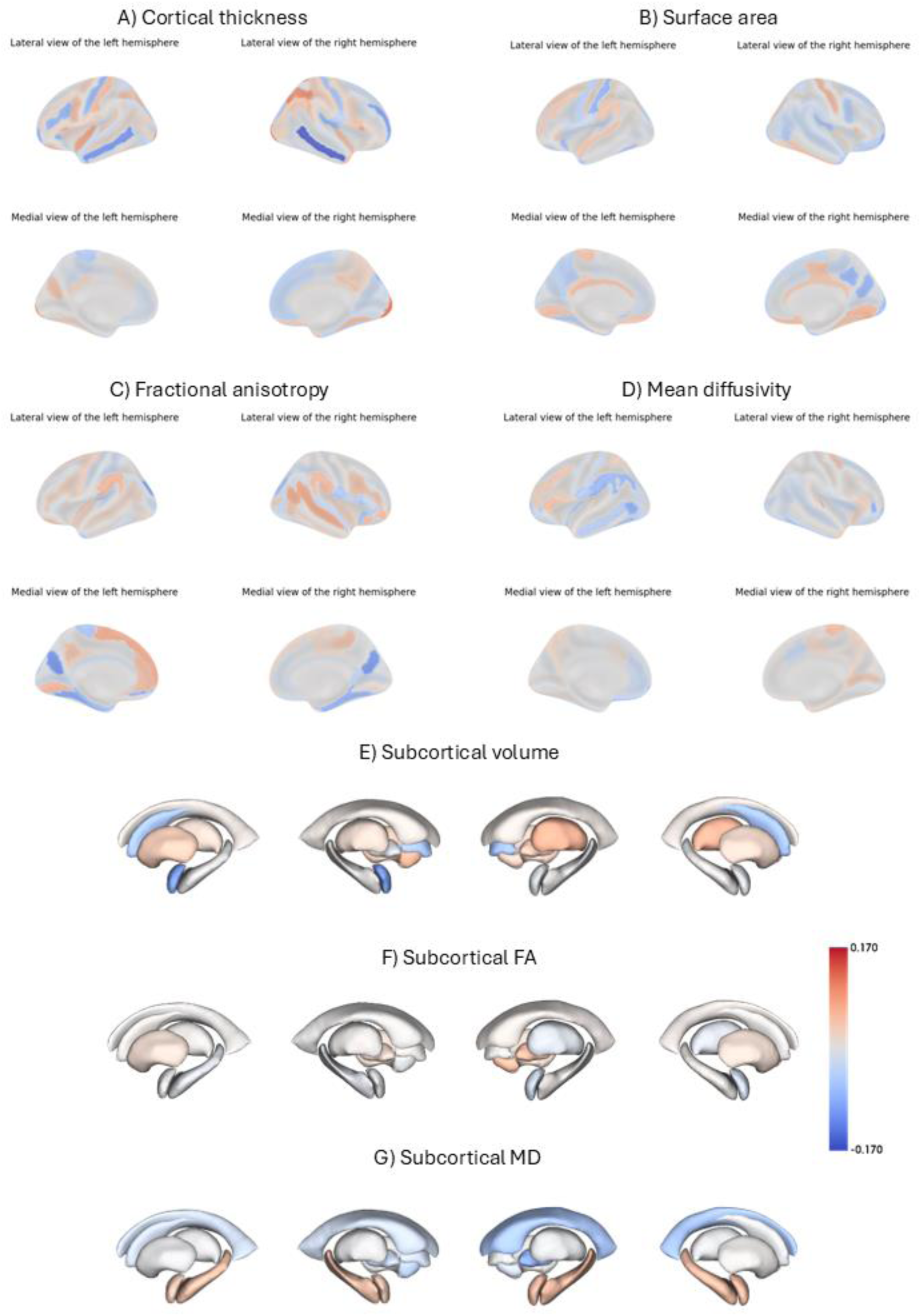
Mean elastic net classification model coefficients for sMRI measures: A) Cortical thickness, B) Cortical surface area, C) White matter fractional anisotropy, D) White matter mean diffusivity, E) Subcortical volume, F) Subcortical fractional anisotropy, G) Subcortical mean diffusivity. Cortical parcellation uses Destrieux atlas and subcortical parcellation uses ASEG subcortical atlas.

In longitudinal weight-trajectory classification models based on structural brain measures and covariates, T1-weighted MRI–derived predictors accounted for 48.6% of total model weight, whereas diffusion tensor imaging–based predictors accounted for 40.8%. Notably, T1-weighted features contributed more strongly to the longitudinal classification task than to the cross-sectional regression task, suggesting that changes in morphometric characteristics may be detectable earlier than alterations in white-matter microstructure in relation to future weight gain. Consistent with the cross-sectional structural MRI models, non-imaging covariates accounted for 10.6% of total model weight, with the polygenic risk score contributing 1.0%, scanner identifiers 5.3%, and sociodemographic background variables 4.2%.

At the level of individual regions of interest and structural features, the largest positive coefficients for classifying participants into weight-gain versus weight-stable groups were associated with cortical thickness of the right occipital pole, cortical thickness of the right intraparietal sulcus and transverse parietal sulci, volume of right cerebellar white matter, cortical thickness of the left occipital pole, and volume of left cerebellar white matter. In contrast, the largest negative coefficients were associated with cortical thickness of the middle temporal gyri bilaterally, fractional anisotropy of white matter adjacent to the parieto-occipital sulci bilaterally, and volume of the left amygdala. Comprehensive lists of mean classification model coefficients and their standard deviations are provided in Online Supplementary Files 5 (imaging data only) and 6 (imaging data with non-imaging covariates).

#### Resting state fMRI longitudinal classification of future weight gain

When predicting weight trajectory—defined as weight gain versus stable weight status at one-year follow-up—from baseline resting-state functional connectivity measures without non-imaging covariates, the model achieved a mean test AUC of 0.60; inclusion of non-imaging covariates increased the mean test AUC to 0.69. Training and test performance metrics for longitudinal classification of weight trajectory are reported in Table 2B. Mean coefficients from the cross-validated elastic net models are visualized as heatmaps of network correlations in Figure 6. Non-imaging covariates are shown in Supplementary Figure S9.

**Figure 6.**
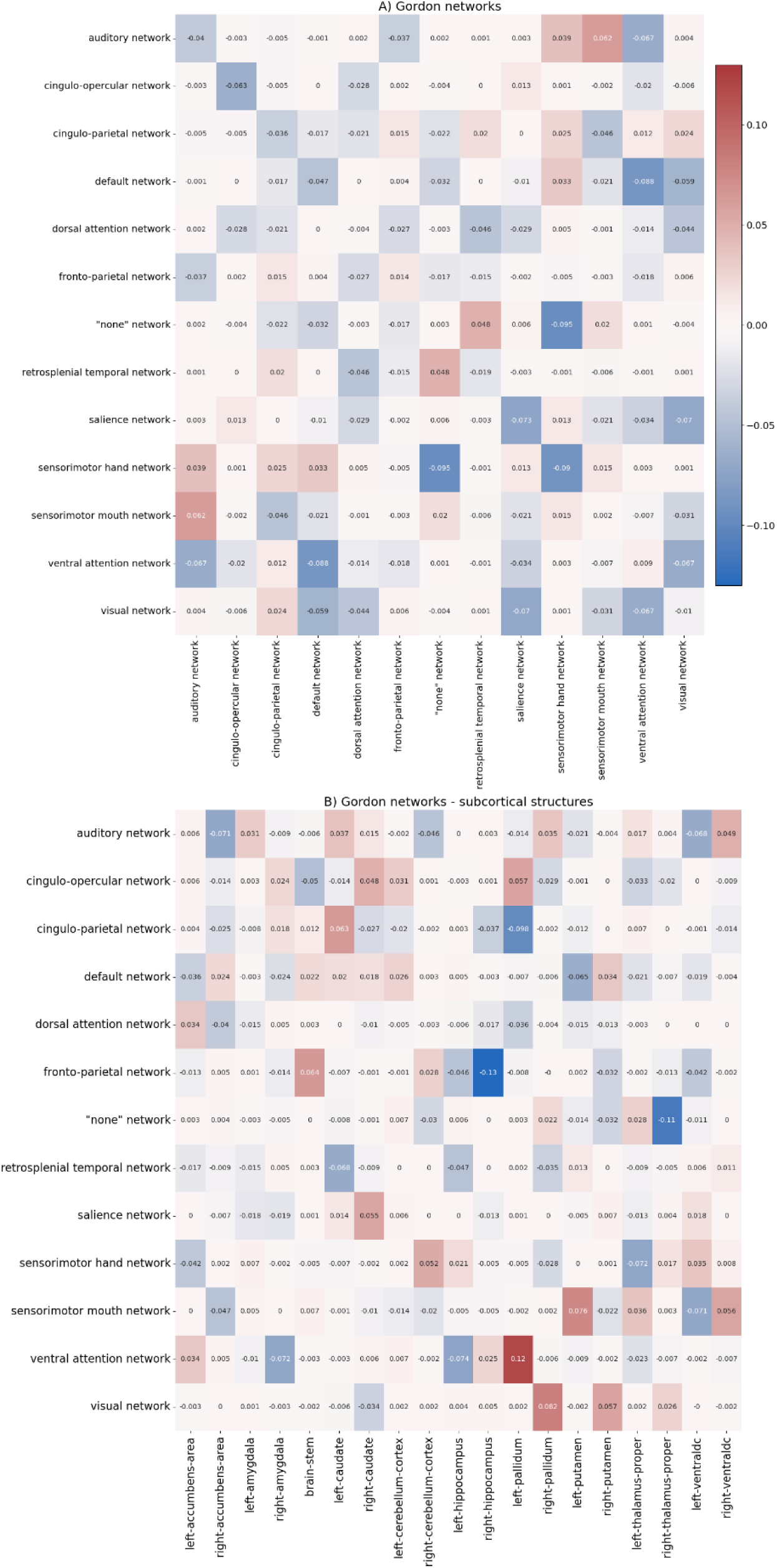
Mean elastic net classification model coefficients for rs-fMRI measures: A) Gordon network intercorrelations, B) Gordon network to subcortex correlations.

Relative contributions of different predictor categories to the aggregate sum of absolute mean coefficients for models including non-imaging covariates are presented in Supplementary Figure S10.

In longitudinal weight-trajectory classification models based on resting-state fMRI measures and covariates, Gordon network intercorrelations accounted for 17.0% of total model weight, whereas correlations between cortical Gordon networks and subcortical regions accounted for 48.2%, with a left-hemisphere predominance (25.8% left vs. 20.9% right). Non-imaging covariates contributed 34.8% of total model weight, with scanner identifiers accounting for 13.9%, sociodemographic background variables for 16.4%, and polygenic risk scores for 4.5%. The relatively larger contribution of non-imaging covariates in these models may reflect the comparatively weaker predictive signal of resting-state functional connectivity features for longitudinal weight-trajectory classification.

At the level of individual connectivity features, the largest positive coefficients were associated with correlations between the ventral attention network and left pallidum, the visual network and right pallidum, the sensorimotor-mouth network and left putamen, the frontoparietal network and brainstem, the cingulo-parietal network and left caudate, and the sensorimotor-mouth and auditory networks. The largest negative coefficients were associated with correlations between the frontoparietal network and right hippocampus, the “none” network (consisting of miscellaneous smaller parcels) and right thalamus-proper, the cingulo-parietal network and left pallidum, the sensorimotor-hand network and the “none” network, and within-network correlations in the sensorimotor-hand network. Comprehensive lists of mean classification model coefficients and their standard deviations are provided in Online Supplementary Files 7 (imaging data only) and 8 (imaging data with non-imaging covariates).

### 3.2 Leave-one-site-out cross-validations

To assess model generalizability across data collection sites, both cross-sectional regression and longitudinal classification analyses were conducted using a leave-one-site-out cross-validation strategy. These analyses were performed separately for structural and resting-state functional brain imaging measures, both with and without the inclusion of non-imaging covariates. Mean test performance metrics from the leave-one-site-out cross-validation analyses are reported in Tables 3A and 3B. Test performance metrics for individual sites are provided in Supplementary Tables S1 and S2. Comprehensive listings of mean model coefficients for all leave-one-site-out analyses are available in Online Supplementary Files 9–16.

### 3.3 Conventional statistics with dimension reduction

Principal component analysis (PCA) was performed separately for each imaging modality to reduce dimensionality and to summarize variance across measures. The number of identified principal components (PCs) and their cumulative explained variance are reported below (see Supplementary Figures S11–S13).

For structural MRI (sMRI) measures (Supplementary Figure S11), four PCs were identified for cortical thickness, accounting for 35.15% of the total variance. Cortical surface area was summarized by three PCs, explaining 40.62% of the variance, while subcortical volume was represented by four PCs, accounting for 64.05% of the variance.

For diffusion tensor imaging (DTI) measures (Supplementary Figure S12), five PCs captured variance in white-matter fractional anisotropy (FA-WM), explaining 67.30% of the total variance. Mean diffusivity in white matter (MD-WM) was summarized by three PCs, accounting for 70.40% of the variance. For subcortical regions, two PCs explained 75.97% of the variance in FA, and four PCs explained 83.26% of the variance in MD.

For resting-state functional MRI (rs-fMRI) measures (Supplementary Figure S13), five PCs summarized subcortical inter-network connectivity (based on the Gordon parcellation), accounting for 35.51% of the variance. Subcortical intra-network connectivity was represented by three PCs, explaining 44.55% of the variance, while cortico-subcortical connectivity was summarized by seven PCs, accounting for 32.46% of the variance.

**Table 4** presents the meta-analysis of neuroimaging-derived components associated with baseline TMI across 21 sites, stratified by sex and scanner type. Overall, baseline TMI showed predominantly negative associations with most neuroimaging components. However, specific components related to subcortical volume (PC3), inter-network connectivity (PC5), and cortico-subcortical network connectivity (PC2) exhibited positive associations with TMI. No significant sex differences were observed (all subgroup p-values > 0.05). Scanner-related differences contributed to elevated heterogeneity in cortical thickness measures.

**Table 4.**
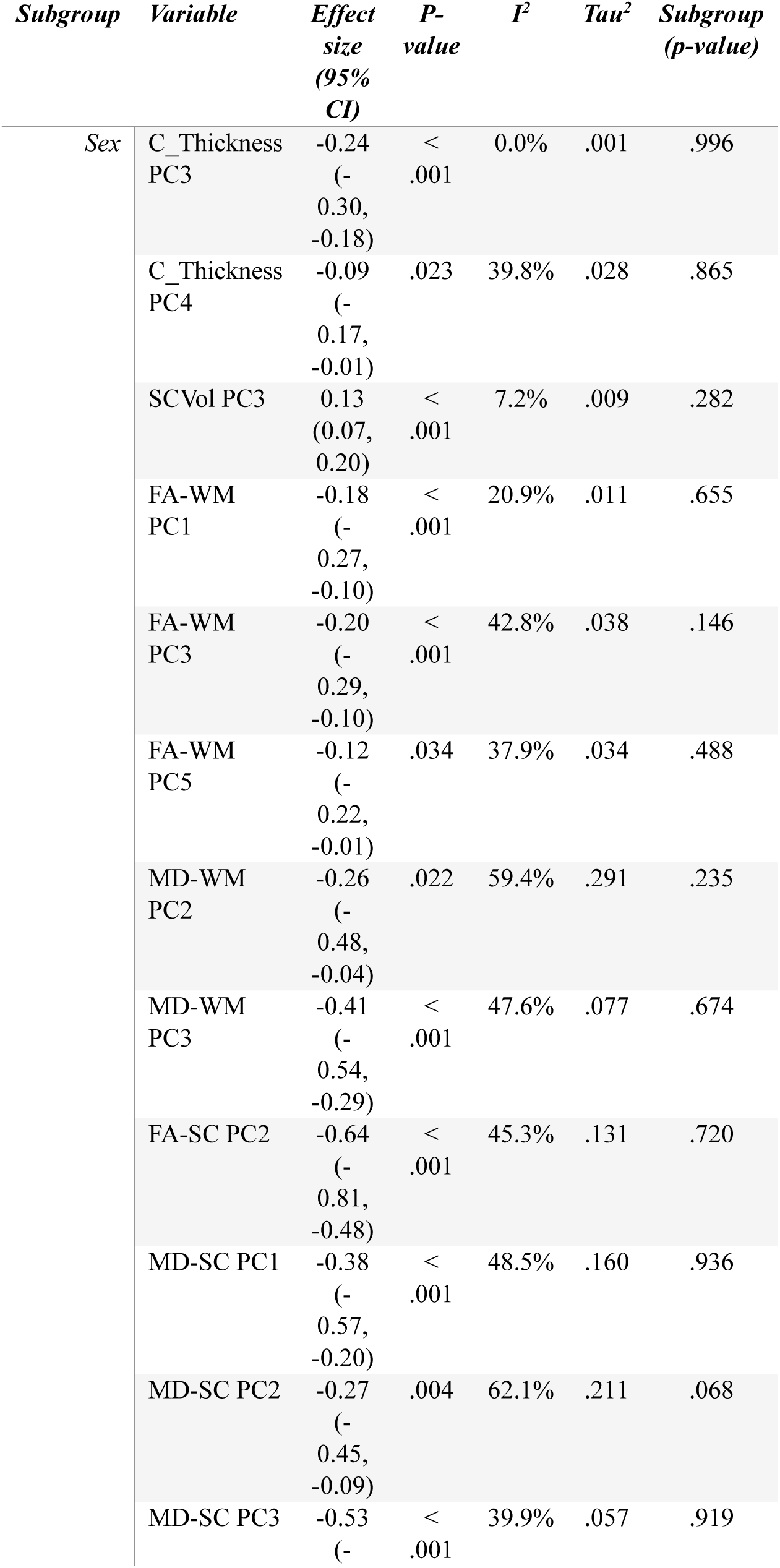
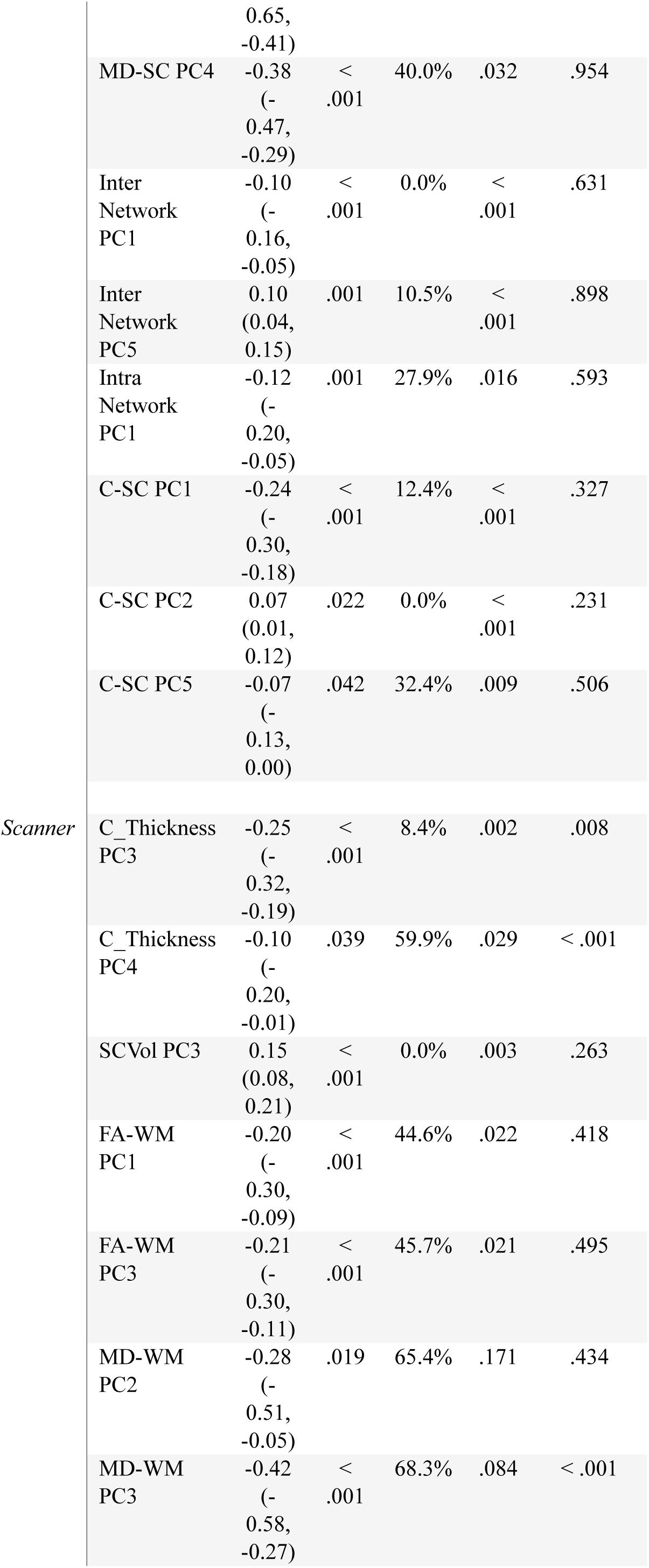
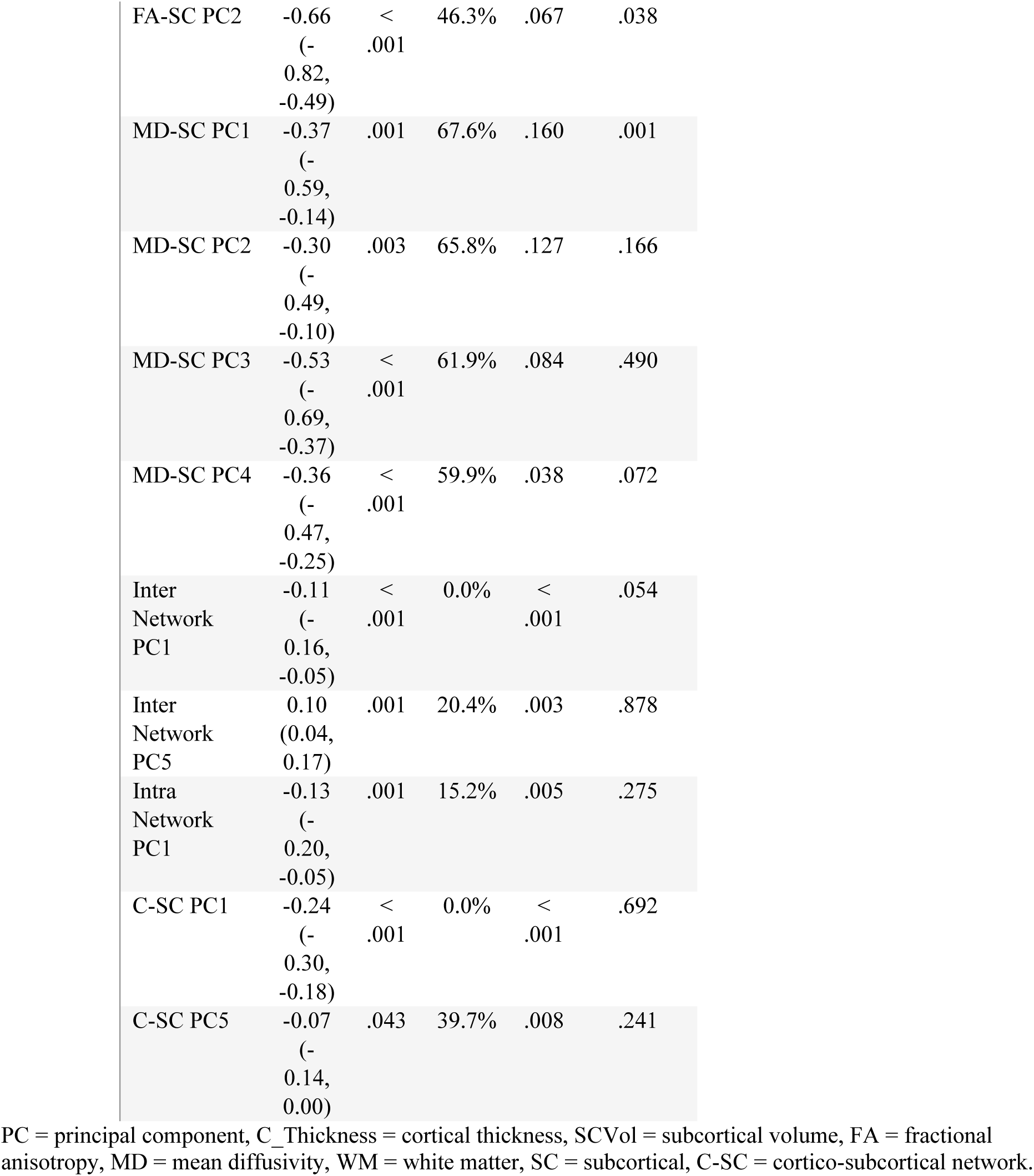
Significant meta-analysis results: neuroimaging components associated with baseline TMI.

Heterogeneity, as indexed by I², was relatively high for diffusivity measures (e.g., MD-WM PC3: 42.8–68.3%; MD-SC PC1: 48.5–67.6%), which may also reflect scanner-related variability. Despite this technical heterogeneity, the directionality of TMI-associated effects—such as cortical thinning and reduced FA—was consistent across scanner subtypes.

**Table 5** presents the meta-analysis of neuroimaging components associated with one-year adiposity change (weight gain vs. weight stability) across sites, stratified by sex and scanner type. Lower cortical thickness (PC3/PC4), reduced subcortical volume (PC4), increased MD-WM (PC2/PC3) and MD-SC (PC3/PC4), as well as greater subcortical volume (PC3), were associated with an elevated risk of weight gain. Alterations in cortico-subcortical connectivity exhibited bidirectional associations: greater intra-network integration (PC1) and cortico-subcortical network connectivity (PC1) were associated with reduced risk, whereas increased inter-network connectivity (PC5) was associated with increased risk of weight gain.

**Table 5.**
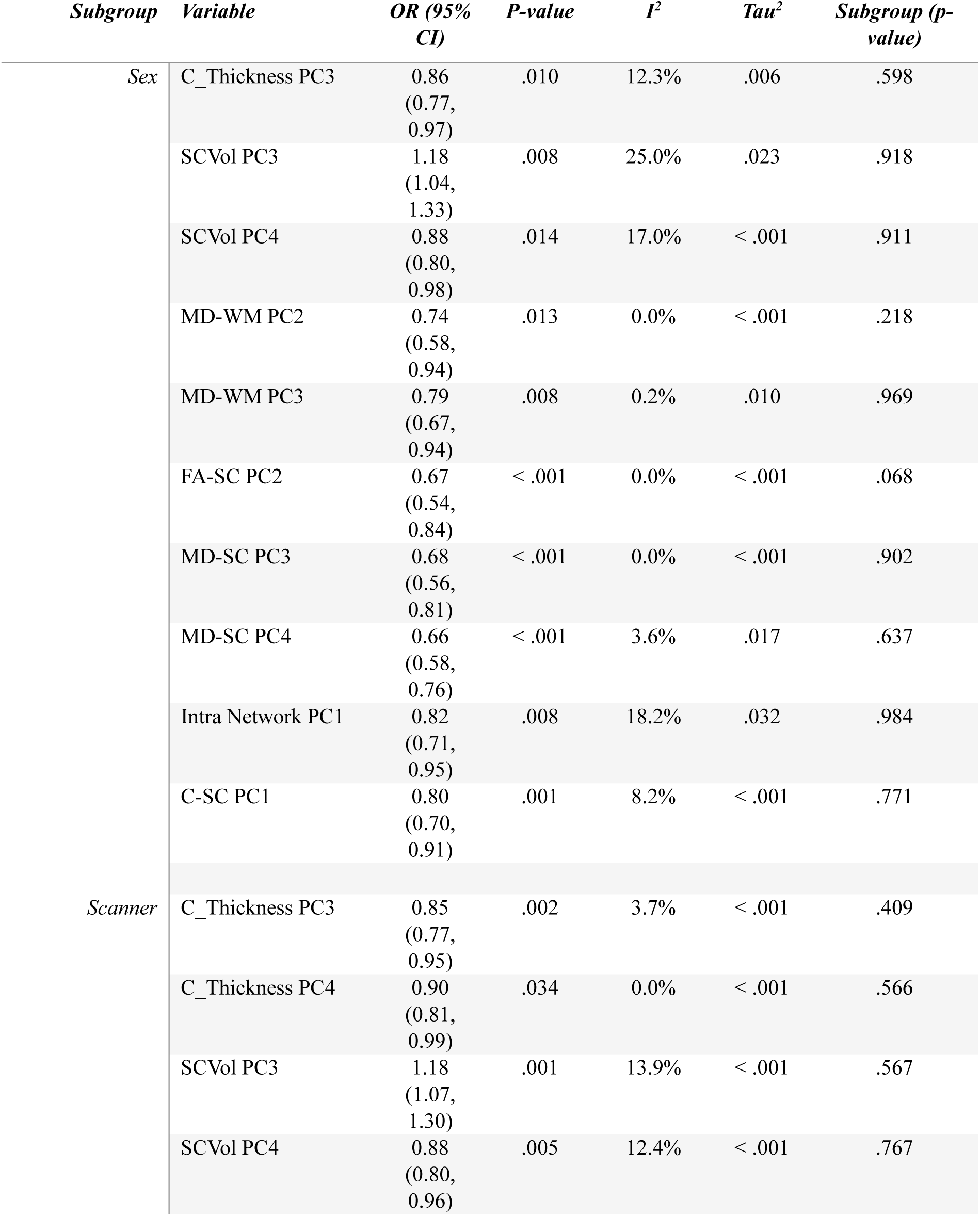
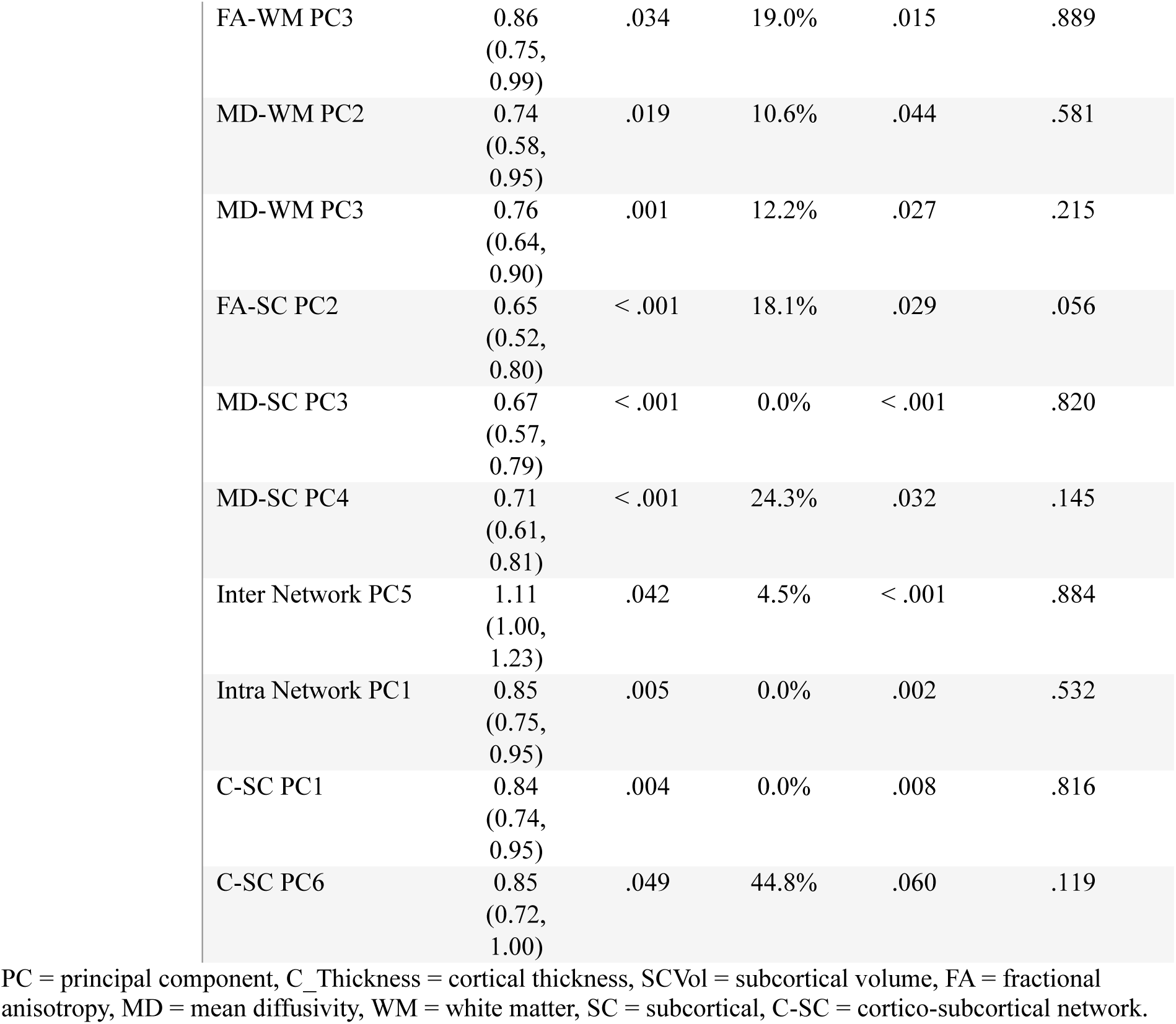
Significant meta-analysis results: neuroimaging components associated with one-year weight gain.

| *Subgroup* | *Variable* | *OR (95% CI)* | *P-value* | *I<sup>2</sup>* | *Tau<sup>2</sup>* | *Subgroup (p-value)* |
| --- | --- | --- | --- | --- | --- | --- |
| *Sex* | C_Thickness PC3 | 0.86<br>(0.77, 0.97) | .010 | 12.3% | .006 | .598 |
|  | SCVol PC3 | 1.18<br>(1.04, 1.33) | .008 | 25.0% | .023 | .918 |
|  | SCVol PC4 | 0.88<br>(0.80, 0.98) | .014 | 17.0% | < .001 | .911 |
|  | MD-WM PC2 | 0.74<br>(0.58, 0.94) | .013 | 0.0% | < .001 | .218 |
|  | MD-WM PC3 | 0.79<br>(0.67, 0.94) | .008 | 0.2% | .010 | .969 |
|  | FA-SC PC2 | 0.67<br>(0.54, 0.84) | < .001 | 0.0% | < .001 | .068 |
|  | MD-SC PC3 | 0.68<br>(0.56, 0.81) | < .001 | 0.0% | < .001 | .902 |
|  | MD-SC PC4 | 0.66<br>(0.58, 0.76) | < .001 | 3.6% | .017 | .637 |
|  | Intra Network PC1 | 0.82<br>(0.71, 0.95) | .008 | 18.2% | .032 | .984 |
|  | C-SC PC1 | 0.80<br>(0.70, 0.91) | .001 | 8.2% | < .001 | .771 |
| *Scanner* | C_Thickness PC3 | 0.85<br>(0.77, 0.95) | .002 | 3.7% | < .001 | .409 |
|  | C_Thickness PC4 | 0.90<br>(0.81, 0.99) | .034 | 0.0% | < .001 | .566 |
|  | SCVol PC3 | 1.18<br>(1.07, 1.30) | .001 | 13.9% | < .001 | .567 |
|  | SCVol PC4 | 0.88<br>(0.80, 0.96) | .005 | 12.4% | < .001 | .767 |

|  |  |  |  |  |  |
| --- | --- | --- | --- | --- | --- |
| FA-WM PC3 | 0.86<br>(0.75,<br>0.99) | .034 | 19.0% | .015 | .889 |
| MD-WM PC2 | 0.74<br>(0.58,<br>0.95) | .019 | 10.6% | .044 | .581 |
| MD-WM PC3 | 0.76<br>(0.64,<br>0.90) | .001 | 12.2% | .027 | .215 |
| FA-SC PC2 | 0.65<br>(0.52,<br>0.80) | < .001 | 18.1% | .029 | .056 |
| MD-SC PC3 | 0.67<br>(0.57,<br>0.79) | < .001 | 0.0% | < .001 | .820 |
| MD-SC PC4 | 0.71<br>(0.61,<br>0.81) | < .001 | 24.3% | .032 | .145 |
| Inter Network PC5 | 1.11<br>(1.00,<br>1.23) | .042 | 4.5% | < .001 | .884 |
| Intra Network PC1 | 0.85<br>(0.75,<br>0.95) | .005 | 0.0% | .002 | .532 |
| C-SC PC1 | 0.84<br>(0.74,<br>0.95) | .004 | 0.0% | .008 | .816 |
| C-SC PC6 | 0.85<br>(0.72,<br>1.00) | .049 | 44.8% | .060 | .119 |

PC = principal component, C\_Thickness = cortical thickness, SCVol = subcortical volume, FA = fractional anisotropy, MD = mean diffusivity, WM = white matter, SC = subcortical, C-SC = cortico-subcortical network.

In this longitudinal framework, overall heterogeneity was low, and no significant sex- or scanner-related differences were observed for any of the reported associations.

## 4 Discussion

In this study, we replicated and extended the analyses and findings of Adise et al.^41^, demonstrating that structural and resting-state fMRI measures predict proportional body weight at baseline and weight gain after one year in a large, ethnically diverse cohort of 9- to 10-year-old children from the ABCD Study®. We leveraged a larger sample size (using the updated ABCD data release 5.0) and relied on a more appropriate index to model proportional weight across development (TMI). In addition, we comprehensively investigated the generalizability of the results using leave-one-site-out cross-validation, included the polygenic risk scores for obesity to address the genetic determinants of obesity, and conducted meta-analytic regression analyses using principal component analysis (PCA) to reduce the dimensionality of the brain imaging data and identify components representing key patterns within the dataset.

Overall, the model trained with structural MRI data (i.e., regional measures of cortical thickness, surface area, subcortical volume, and FA and MD diffusion metrics) outperformed the model trained with functional MRI data. This superior performance may be attributable to the inclusion of both T1- and DTI-based measures within the structural modality, which together provide a broader range of complementary information relevant to predicting body adiposity. Moreover, the stronger performance of structural MRI data compared with functional MRI data may reflect that structural measures more directly capture neuroanatomical differences associated with elevated body weight and changes in weight.

### Comparison of current results to literature

#### Cortical thickness / surface area

Prior literature has reported both negative and positive associations between high body fat and cortical thickness (CT) in children and adolescents in multiple cortical regions^55–57^, and particularly in the prefrontal cortex^8,25,58–60^. Similarly, both negative and positive correlations between cortical surface area and body fat have been reported in various regions across the cortex^55,61^. Consistent with previous literature, we observed widespread associations of CT and surface area with TMI in our data-analysis. In particular, we observed a similar regional pattern of reduced cortical thickness as reported in prior literature^8^, although largely localized in the right hemisphere. The negative association with cortical thickness is more prominent in the cross-sectional regression model as compared to the longitudinal classification model, consistent with the notion of cortical thinning in obesity.^62^

#### Subcortical volume

Previously reported associations between volumes of subcortical structures and high body fat include negative correlations in caudate^63,64^, putamen^64,65^, left hippocampus^26,66,67^ and cerebellum bilaterally^68^, and positive correlations in globus pallidus^27,66,69^ and nucleus accumbens^13,70,71^. Findings in amygdala^13,64^ and right hippocampus^64,72–74^ have been mixed. Additionally, Adise et al. (2024) report negative associations between body fat and volumes of various subcortical structures in preadolescent girls but not boys^75^. We observed the negative associations to caudate volume bilaterally, with stronger negative association in the longitudinal classification model, suggesting that reduction in caudate volume may precede weight gain. We also observed a negative association in left hippocampus in the cross-sectional regression model, but not the longitudinal classification model, which may suggest that the reduction of volume in left hippocampus occurs following weight gain. This notion is broadly consistent with a sequential pattern of subcortical volume reductions reported by Zhou et al.^76^ and interpreted within a directional network model. We additionally observed both negative and positive associations between body adiposity and the volume of reward-related subcortical structures. Cerebellar volume, a structure implicated in reward anticipation and processing^77^, showed negative associations bilaterally. Globus pallidus volume showed positive associations bilaterally in cross-sectional analyses and in the right hemisphere in longitudinal classification. Nucleus accumbens volume was positively associated with future weight gain only in the longitudinal model, suggesting that increased volume of nucleus accumbens may be related to future weight gain. Nucleus accumbens is considered a hotspot for hedonic “wanting” reaction,^78^ possibly explaining its association specifically with future weight gain.

#### Subcortical DTI measures

Prior diffusion weighted imaging research focusing on subcortical structures is scarce, and seems to favor advanced diffusion imaging techniques, such as restricted spectrum imaging and diffusion basis spectrum imaging, and prior results are therefore not directly comparable to the present findings. In brief, prior studies have found associations of childhood BMI, obesity or waist circumference and RSI or DBSI measurements in nucleus accumbens^14,79,80^, caudate^79,80^, pallidum^79^, putamen^79,80^, thalamus^79^, ventral diencephalon^79^, right amygdala^79^ and hypothalamus^80^. Current results, while not directly comparable, show associations with DTI measures in nucleus accumbens (decreased MD and increased FA), caudate (increased MD) and putamen (increased MD and FA) of the striatum as well as amygdala (increased MD) in the cross-sectional regression model. The model coefficients associated with subcortical DTI measures were smaller in the longitudinal classification model, possibly suggesting that subcortical microstructural alterations are more closely associated with concurrent adiposity than with subsequent weight gain.

#### fMRI

In prior research, both positive and negative associations between high body fat and connectivity of various resting-state functional networks have been reported. Negative associations have been found for connections within salience network^81^ and cingulo-opercular network^81^ and between salience network and other networks such as cingulo-opercular^81^ and default mode network^82^, as well as amygdala^82^. Positive associations have also been found within salience network^82^ and between sensorimotor hand and visual, as well as default mode and dorsal attention networks^81^. Kaltenhauser et al. note that both salience network and cingulo-opercular network - both negatively associated with high BMI - encompass the dorsal anterior cingulate cortex, which is involved in cognitive control, motivation and reward-based decision making.^81^ Current results are again in partial agreement with prior findings, e.g., concerning the negative associations within salience and cingulo-opercular networks in the cross-sectional regression model, possibly reflecting altered engagement of cognitive control and reward-related networks. In general, correlations involving regions of the reward system, including striatal structures and (e.g.,) the retrosplenial temporal network had large positive coefficients. Notably, we also observed a positive association for correlation between dorsal attention network and left accumbens area, previously linked to obesity and thought to relate to a reorganization of brain connectivity towards orienting to external stimuli.^83^ Finally, current results show negative associations involving cingulo-opercular and fronto-parietal networks, both implicated in self-regulation system, important for regulating appetitive behavior.^84^

#### DTI

Associations between BMI, overweight or obesity and white matter integrity in children as measured by DTI reported in prior literature have been largely negative. Global negative associations have been reported^85,86^, as well as local negative associations in canonical white matter tracts, including corpus callosum and fornix^87,88^, cingulum and corona radiata^87^, left superior and inferior longitudinal fasciculi^89^, bilateral uncinate fasciculi and left inferior fronto-occipital fasciculus^15^ as well as left temporal stem, bilateral optic radiation, left internal and external capsule and left splenium^90^. Some studies have also reported local positive associations between obesity and white matter integrity in left association and projection fibers^91^, right interior fronto-occipital fasciculus and left cingulum^15^, as well as in right prefrontal region with partial gray matter contamination^90^. While most of the prior research investigates associations of microstructure of white matter tracts and BMI or obesity, the current analysis was based on cortical parcellation and therefore cannot be directly compared.

#### Methodological considerations

The impact of puberty on proportional body mass is important to consider in the age range of the current study, as BMI starts to rise before puberty and continues to rise through adolescence.^92^ The association of the level of pubertal development and BMI is further complicated by the complex and sex-dependent interaction between BMI and the time of onset of puberty. In brief, overweight and obesity are associated with earlier onset of puberty in girls, whereas in boys, obesity (but not overweight) is associated with later onset of puberty.^93^ Accordingly, pubertal developmental index was among the most powerful predictors of TMI in our analysis.

While there is evidence of negative associations between BMI and CT across the lifespan, particularly in the fronto-occipital cortical regions^94^, some evidence also exists linking obesity and high body mass with increased CT, as well as of no association to either direction^95^, suggesting a more complex relation between body adiposity and CT. Gomez-apo et al. ^96^ note that the inconsistency in some of the reported associations between body mass and brain structural measures may result from sample differences as well as different imaging and preprocessing parameters. Importantly for the present study, Sharkey et al. found no association between cortical thickness and BMI in adolescent participants, concluding that cortical thinning related to high body adiposity develops after adolescence.^97^ In brief, differences in sample characteristics, imaging acquisition and processing parameters, as well as the developmental stage of the participants may influence observed associations and should be considered when interpreting the results.

Between-site variability presents a challenge for machine learning in multi-site neuroimaging studies, as demonstrated in the present analyses, as the variability in data distribution between sites can negatively impact the model performance on the combined multi-site dataset. This effect can be mitigated with the use of harmonization algorithms^98^, although there are some reports of data harmonization not improving or even degrading the performance^99,100^.

Regardless of the effect on performance, current harmonization algorithms require data from all study sites to be available at model training, for which reason the methodology is not suitable for harmonization of training datasets for the purpose of generalization of completely novel sites. Further, regular “pooled” cross-validation methods, such as k-fold cross-validation with random partition do not estimate model generalization to completely novel site-specific datasets in a multi-site setting, as the randomness of the partition implies that the model is able to sample from all sites present in the pooled dataset. For these reasons, we performed leave-one-site-out cross-validation to estimate model generalization to such novel site-specific datasets not available for the model to sample at training. We observed high variance between test-sites and conclude that traditional machine learning models such as elastic net may not generalize reliably to out-of-sample sites in the presence of substantial site-related variability.

### Limitations

Although the ABCD tabular data release contains an extensive selection of both imaging and non-imaging measurements for the large cohort of participants, it does not contain a parcellation of the hypothalamus, a key region in energy homeostasis whose neuroinflammation may contribute to the development of obesity. In addition, the present study does not consider measures of microstructural integrity of canonical white matter tracts. Consequently, this study does not include all measurements from brain regions known from prior literature to be involved in weight gain and the development of obesity and may therefore underestimate the associations of high body adiposity and brain structure and function.

### Conclusions

Our results demonstrate associations between brain-derived measures and proportional body weight in late childhood and future weight gain, in both machine learning and traditional statistical frameworks. Further, our results highlight the challenge posed by between-site variability for machine learning model generalization across datasets in machine learning neuroimaging research, underscoring the need for careful validation strategies and methodological advances in multi-site neuroimaging research.

### Data availability statement

The data used in this study were drawn from the Adolescent Brain Cognitive Development (ABCD) Study® Curated Annual Release 5.0 (DOI: *10.15154/8873-zj65*). The ABCD Study® is a large-scale, longitudinal project supported by multiple NIH institutes, designed to track brain, cognitive, and behavioral development in nearly 12,000 U.S. youth.

### Code availability

The code used in the machine learning data-analysis presented in this study will be made available upon acceptance of this manuscript.

### Funding

**This study was funded by grants to JJT:**

**Finnish Medical Foundation;**

**Emil Aaltonen Foundation;**

**Sigrid Jusélius Foundation;**

**Hospital District of Southwest Finland, Finnish State Grants for Clinical Research (VTR);**

**Signe and Ane Gyllenberg Foundation;**

**Open Access publication funding was provided by University of Turku (including Turku University Central Hospital).**

**EPP was supported by Turku University Foundation**

**NH was funded by the Jenny and Antti Wihuri Foundation and the Hospital District of Southwest Finland, Finnish State Grants for Clinical Research (VTR).**

**HKA was supported by University of Turku Graduate School, and Yrjö Jahnsson Foundation**

**RL was funded by the Yrjö Jahnsson Foundation.**

### Author contributions

J.J.T. conceived the project. I.S. designed and implemented the machine learning data analysis and was the main contributor to writing the manuscript, with input from all authors R.L. implemented the statistical meta-analysis and made significant contributions to writing the manuscript. MK, RK, RL and EP computed the polygenic risk scores used in the data-analysis.

## Acknowledgements

We thank the ABCD consortium investigators, participating families, and all study staff for their invaluable contributions. ABCD investigators designed and implemented the study; however, they did not participate in the analysis or writing of this report. The opinions and conclusions expressed here are solely those of the authors and do not reflect the official views of the ABCD Study® or its federal sponsors.

## References

1. Mohajan D, Mohajan HK. Obesity and Its Related Diseases: A New Escalating Alarming in Global Health. J Innov Med Res. 2023;2(3):12–23.

2. Obesity and overweight. Accessed October 9, 2025. https://www.who.int/news-room/fact-sheets/detail/obesity-and-overweight

3. Kosti RI, Panagiotakos DB. The epidemic of obesity in children and adolescents in the world. Cent Eur J Public Health. 2006;14(4):151–159. doi:10.21101/cejph.a3398

4. Halfon N, Larson K, Slusser W. Associations between obesity and comorbid mental health, developmental, and physical health conditions in a nationally representative sample of US children aged 10 to 17. Acad Pediatr. 2013;13(1):6–13. doi:10.1016/j.acap.2012.10.007

5. Schwartz MW, Seeley RJ, Zeltser LM, et al. Obesity Pathogenesis: An Endocrine Society Scientific Statement. Endocr Rev. 2017;38(4):267–296. doi:10.1210/er.2017-00111

6. Yu YH. Making sense of metabolic obesity and hedonic obesity. J Diabetes. 2017;9(7):656–666. doi:10.1111/1753-0407.12529

7. Duis J, Butler MG. Syndromic and Nonsyndromic Obesity: Underlying Genetic Causes in Humans. Adv Biol. 2022;6(10):2101154. doi:10.1002/adbi.202101154

8. Laurent JS, Watts R, Adise S, et al. Associations Among Body Mass Index, Cortical Thickness, and Executive Function in Children. JAMA Pediatr. 2020;174(2):170–177. doi:10.1001/jamapediatrics.2019.4708

9. Mestre ZL, Bischoff-Grethe A, Eichen DM, Wierenga CE, Strong D, Boutelle KN. Hippocampal atrophy and altered brain responses to pleasant tastes among obese compared with healthy weight children. Int J Obes 2005. 2017;41(10):1496–1502. doi:10.1038/ijo.2017.130

10. de Groot CJ, van den Akker ELT, Rings EHHM, Delemarre-van de Waal HA, van der Grond J. Brain structure, executive function and appetitive traits in adolescent obesity. Pediatr Obes. 2017;12(4):e33–e36. doi:10.1111/ijpo.12149

11. Alosco ML, Stanek KM, Galioto R, et al. Body mass index and brain structure in healthy children and adolescents. Int J Neurosci. 2014;124(1):49–55. doi:10.3109/00207454.2013.817408

12. Ronan L, Alexander-Bloch A, Fletcher PC. Childhood Obesity, Cortical Structure, and Executive Function in Healthy Children. Cereb Cortex N Y N 1991. 2020;30(4):2519–2528. doi:10.1093/cercor/bhz257

13. Perlaki G, Molnar D, Smeets PAM, et al. Volumetric gray matter measures of amygdala and accumbens in childhood overweight/obesity. PloS One. 2018;13(10):e0205331. doi:10.1371/journal.pone.0205331

14. Rapuano KM, Zieselman AL, Kelley WM, Sargent JD, Heatherton TF, Gilbert-Diamond D. Genetic risk for obesity predicts nucleus accumbens size and responsivity to real-world food cues. Proc Natl Acad Sci. 2017;114(1):160–165. doi:10.1073/pnas.1605548113

15. Carbine KA, Duraccio KM, Hedges-Muncy A, Barnett KA, Kirwan CB, Jensen CD. White matter integrity disparities between normal-weight and overweight/obese adolescents: an automated fiber quantification tractography study. Brain Imaging Behav. 2020;14(1):308–319. doi:10.1007/s11682-019-00036-4

16. Black WR, Lepping RJ, Bruce AS, et al. Tonic hyper-connectivity of reward neurocircuitry in obese children. Obes Silver Spring Md. 2014;22(7):1590–1593. doi:10.1002/oby.20741

17. Moreno-Lopez L, Contreras-Rodriguez O, Soriano-Mas C, Stamatakis EA, Verdejo-Garcia A. Disrupted functional connectivity in adolescent obesity. NeuroImage Clin. 2016;12:262–268. doi:10.1016/j.nicl.2016.07.005

18. Yokum S, Gearhardt AN, Harris JL, Brownell KD, Stice E. Individual differences in striatum activity to food commercials predict weight gain in adolescents. Obes Silver Spring Md. 2014;22(12):2544–2551. doi:10.1002/oby.20882

19. Bohon C. Brain response to taste in overweight children: A pilot feasibility study. PloS One. 2017;12(2):e0172604. doi:10.1371/journal.pone.0172604

20. Batterink L, Yokum S, Stice E. Body mass correlates inversely with inhibitory control in response to food among adolescent girls: an fMRI study. NeuroImage. 2010;52(4):1696–1703. doi:10.1016/j.neuroimage.2010.05.059

21. Bruce AS, Holsen LM, Chambers RJ, et al. Obese children show hyperactivation to food pictures in brain networks linked to motivation, reward and cognitive control. Int J Obes 2005. 2010;34(10):1494–1500. doi:10.1038/ijo.2010.84

22. Stice E, Yokum S. Gain in Body Fat Is Associated with Increased Striatal Response to Palatable Food Cues, whereas Body Fat Stability Is Associated with Decreased Striatal Response. J Neurosci. 2016;36(26):6949–6956. doi:10.1523/JNEUROSCI.4365-15.2016

23. Davids S, Lauffer H, Thoms K, et al. Increased dorsolateral prefrontal cortex activation in obese children during observation of food stimuli. Int J Obes 2005. 2010;34(1):94–104. doi:10.1038/ijo.2009.193

24. Bruce AS, Lepping RJ, Bruce JM, et al. Brain responses to food logos in obese and healthy weight children. J Pediatr. 2013;162(4):759–764.e2. doi:10.1016/j.jpeds.2012.10.003

25. Ronan L, Alexander-Bloch A, Fletcher PC. Childhood Obesity, Cortical Structure, and Executive Function in Healthy Children. Cereb Cortex N Y N 1991. 2020;30(4):2519–2528.doi:10.1093/cercor/bhz257

26. Mestre ZL, Bischoff-Grethe A, Eichen DM, Wierenga CE, Strong D, Boutelle KN. Hippocampal atrophy and altered brain responses to pleasant tastes among obese compared with healthy weight children. Int J Obes 2005. 2017;41 (10):1496–1502. doi:10.1038/ijo.2017.130

27. de Groot CJ, van den Akker ELT, Rings EHHM, Delemarre-van de Waal HA, van der Grond J. Brain structure, executive function and appetitive traits in adolescent obesity. Pediatr Obes. 2017;12(4):e33–e36. doi:10.1111/ijpo.12149

28. Sharkey RJ, Karama S, Dagher A. Overweight is not associated with cortical thickness alterations in children. Front Neurosci. 2015;9:24. doi:10.3389/fnins.2015.00024

29. Boutelle KN, Wierenga CE, Bischoff-Grethe A, et al. Increased brain response to appetitive tastes in the insula and amygdala in obese compared with healthy weight children when sated. Int J Obes 2005. 2015;39(4):620–628. doi:10.1038/ijo.2014.206

30. Adise S, Geier CF, Roberts NJ, White CN, Keller KL. Is brain response to food rewards related to overeating? A test of the reward surfeit model of overeating in children. Appetite. 2018;128:167–179. doi:10.1016/j.appet.2018.06.014

31. Adise S, Geier CF, Roberts NJ, White CN, Keller KL. Food or money? Children’s brains respond differently to rewards regardless of weight status. Pediatr Obes. 2019;14(2):e12469. doi:10.1111/ijpo.12469

32. Tomiyama AJ, Carr D, Granberg EM, et al. How and why weight stigma drives the obesity “epidemic” and harms health. BMC Med. 2018;16(1):123. doi:10.1186/s12916-018-1116-5

33. Yang J, Manolio TA, Pasquale LR, et al. Genome partitioning of genetic variation for complex traits using common SNPs. Nat Genet. 2011;43(6):519–525. doi:10.1038/ng.823

34. Lewis CM, Vassos E. Polygenic risk scores: from research tools to clinical instruments. Genome Med. 2020;12(1):44. doi:10.1186/s13073-020-00742-5

35. Khera AV, Chaffin M, Wade KH, et al. Polygenic Prediction of Weight and Obesity Trajectories from Birth to Adulthood. Cell. 2019;177(3):587–596.e9. doi:10.1016/j.cell.2019.03.028

36. Peterson CM, Su H, Thomas DM, et al. Tri-Ponderal Mass Index vs Body Mass Index in Estimating Body Fat During Adolescence. JAMA Pediatr. 2017;171(7):629–636. doi:10.1001/jamapediatrics.2017.0460

37. Özyildirim C, Unsal EN, Ayhan NY. Performance of triponderal mass index, body mass index z scores, and body mass index performance in the diagnosis of obesity in children and adolescents. Nutr Burbank Los Angel Cty Calif. 2023;114:112116. doi:10.1016/j.nut.2023.112116

38. Ued F, Castro MJS, Bardi LR, et al. Triponderal Mass Index rather than Body Mass Index in discriminating high adiposity in Brazilian children and adolescents. Nutr Hosp. Published online November 5, 2024. doi:10.20960/nh.05432

39. De Lorenzo A, Romano L, Di Renzo L, et al. Triponderal mass index rather than body mass index: An indicator of high adiposity in Italian children and adolescents. Nutr Burbank Los Angel Cty Calif. 2019;60:41–47. doi:10.1016/j.nut.2018.09.007

40. Gul Siraz U, Hatipoglu N, Mazicioglu MM, Ozturk A, Cicek B, Kurtoglu S. Triponderal mass index is as strong as body mass index in the determination of obesity and adiposity. Nutr Burbank Los Angel Cty Calif. 2023;105:111846. doi:10.1016/j.nut.2022.111846

41. Adise S, Allgaier N, Laurent J, et al. Multimodal brain predictors of current weight and weight gain in children enrolled in the ABCD study ®. Dev Cogn Neurosci. 2021;49:100948. doi:10.1016/j.dcn.2021.100948

42. Volkow ND, Koob GF, Croyle RT, et al. The conception of the ABCD study: From substance use to a broad NIH collaboration. Dev Cogn Neurosci. 2018;32:4–7. doi:10.1016/j.dcn.2017.10.002

43. Smith DM, Loughnan R, Friedman NP, et al. Heritability Estimation of Cognitive Phenotypes in the ABCD Study® Using Mixed Models. Behav Genet. 2023;53(3):169–188. doi:10.1007/s10519-023-10141-2

44. Destrieux C, Fischl B, Dale A, Halgren E. Automatic parcellation of human cortical gyri and sulci using standard anatomical nomenclature. NeuroImage. 2010;53(1):1–15. doi:10.1016/j.neuroimage.2010.06.010

45. Fischl B, Salat DH, Busa E, et al. Whole brain segmentation: automated labeling of neuroanatomical structures in the human brain. Neuron. 2002;33(3):341–355. doi:10.1016/s0896-6273(02)00569-x

46. Gordon EM, Laumann TO, Adeyemo B, Huckins JF, Kelley WM, Petersen SE. Generation and Evaluation of a Cortical Area Parcellation from Resting-State Correlations. Cereb Cortex. 2016;26(1):288–303. doi:10.1093/cercor/bhu239

47. Hagler DJ, Hatton S, Cornejo MD, et al. Image processing and analysis methods for the Adolescent Brain Cognitive Development Study. NeuroImage. 2019;202:116091. doi:10.1016/j.neuroimage.2019.116091

48. Geras K, Sutton C. Multiple-source cross-validation. In: Proceedings of the 30th International Conference on Machine Learning. PMLR; 2013:1292-1300. Accessed June 25, 2026. https://proceedings.mlr.press/v28/geras13.html

49. Knight J, Taylor GW, Khademi A. Voxel-Wise Logistic Regression and Leave-One-Source-Out Cross Validation for white matter hyperintensity segmentation. Magn Reson Imaging. 2018;54:119–136. doi:10.1016/j.mri.2018.06.009

50. Leinonen T, Wong D, Vasankari A, et al. Empirical investigation of multi-source cross-validation in clinical ECG classification. Comput Biol Med. 2024;183:109271. doi:10.1016/j.compbiomed.2024.109271

51. Rossum GV, Drake FL. The Python Language Reference Manual. Network Theory Limited; 2011.

52. Harris CR, Millman KJ, van der Walt SJ, et al. Array programming with NumPy. Nature. 2020;585(7825):7825. doi:10.1038/s41586-020-2649-2

53. McKinney W. Data Structures for Statistical Computing in Python. In: 2010:56-61. doi:10.25080/Majora-92bf1922-00a

54. Pedregosa F, Varoquaux G, Gramfort A, et al. Scikit-learn: Machine Learning in Python. J Mach Learn Res. 2011;12(85):2825–2830.

55. Kulisch LK, Arumäe K, Briley DA, Vainik U. Triangulating causality between childhood obesity and neurobehavior: Behavioral genetic and longitudinal evidence. Dev Sci. 2023;26(6):e13392. doi:10.1111/desc.13392

56. Rajan L, McKay CC, Santos Malavé G, et al. Effects of severe obesity and sleeve gastrectomy on cortical thickness in adolescents. Obes Silver Spring Md. 2021;29(9):1516–1525. doi:10.1002/oby.23206

57. Steegers C, Blok E, Lamballais S, et al. The association between body mass index and brain morphology in children: a population-based study. Brain Struct Funct. 2021;226(3):787–800. doi:10.1007/s00429-020-02209-0

58. Kim MS, Luo S, Azad A, et al. Prefrontal Cortex and Amygdala Subregion Morphology Are Associated With Obesity and Dietary Self-control in Children and Adolescents. Front Hum Neurosci. 2020;14:563415. doi:10.3389/fnhum.2020.563415

59. Hall PA, Best JR, Beaton EA, Sakib MN, Danckert J. Morphology of the prefrontal cortex predicts body composition in early adolescence: cognitive mediators and environmental moderators in the ABCD Study. Soc Cogn Affect Neurosci. 2023;18(1):nsab104. doi:10.1093/scan/nsab104

60. Cui J, Li G, Zhang M, et al. Associations between body mass index, sleep-disordered breathing, brain structure, and behavior in healthy children. Cereb Cortex N Y N 1991. 2023;33(18):10087–10097. doi:10.1093/cercor/bhad267

61. Prats-Soteras X, Jurado MA, Ottino-González J, et al. Inflammatory agents partially explain associations between cortical thickness, surface area, and body mass in adolescents and young adulthood. Int J Obes 2005. 2020;44(7):1487–1496. doi:10.1038/s41366-020-0582-y

62. Marqués-Iturria I, Pueyo R, Garolera M, et al. Frontal cortical thinning and subcortical volume reductions in early adulthood obesity. Psychiatry Res Neuroimaging. 2013;214(2):109–115. doi:10.1016/j.pscychresns.2013.06.004

63. Kennedy JT, Collins PF, Luciana M. Higher Adolescent Body Mass Index Is Associated with Lower Regional Gray and White Matter Volumes and Lower Levels of Positive Emotionality. Front Neurosci. 2016;10:413. doi:10.3389/fnins.2016.00413

64. Nouwen A, Chambers A, Chechlacz M, et al. Microstructural abnormalities in white and gray matter in obese adolescents with and without type 2 diabetes. NeuroImage Clin. 2017;16:43–51. doi:10.1016/j.nicl.2017.07.004

65. Yokum S, Stice E. Initial body fat gain is related to brain volume changes in adolescents: A repeated-measures voxel-based morphometry study. Obes Silver Spring Md. 2017;25(2):401–407. doi:10.1002/oby.21728

66. Bauer CCC, Moreno B, González-Santos L, Concha L, Barquera S, Barrios FA. Child overweight and obesity are associated with reduced executive cognitive performance and brain alterations: a magnetic resonance imaging study in Mexican children. Pediatr Obes. 2015;10(3):196–204. doi:10.1111/ijpo.241

67. Lynch KM, Page KA, Shi Y, Xiang AH, Toga AW, Clark KA. The effect of body mass index on hippocampal morphology and memory performance in late childhood and adolescence. Hippocampus. 2021;31(2):189–200. doi:10.1002/hipo.23280

68. García-García I, Michaud A, Dadar M, et al. Neuroanatomical differences in obesity: meta-analytic findings and their validation in an independent dataset. Int J Obes. 2019;43(5):943–951. doi:10.1038/s41366-018-0164-4

69. Zaugg KK, Cobia DJ, Jensen CD. Abnormalities in deep-brain morphology and orbitofrontal cortical thinning relate to reward processing and body mass in adolescent girls. Int J Obes 2005. 2022;46(9):1720–1727. doi:10.1038/s41366-022-01188-y

70. García-García I, Morys F, Dagher A. Nucleus accumbens volume is related to obesity measures in an age-dependent fashion. J Neuroendocrinol. 2020;32(12):e12812. doi:10.1111/jne.12812

71. Kakoschke N, Lorenzetti V, Caeyenberghs K, Verdejo-García A. Impulsivity and body fat accumulation are linked to cortical and subcortical brain volumes among adolescents and adults. Sci Rep. 2019;9(1):2580. doi:10.1038/s41598-019-38846-7

72. Masterson TD, Bobak C, Rapuano KM, Shearrer GE, Gilbert-Diamond D. Association between regional brain volumes and BMI z-score change over one year in children. PloS One. 2019;14(9):e0221995. doi:10.1371/journal.pone.0221995

73. Hashimoto T, Takeuchi H, Taki Y, et al. Increased posterior hippocampal volumes in children with lower increase in body mass index: a 3-year longitudinal MRI study. Dev Neurosci. 2015;37(2):153–160. doi:10.1159/000370064

74. Moreno-López L, Soriano-Mas C, Delgado-Rico E, Rio-Valle JS, Verdejo-García A. Brain structural correlates of reward sensitivity and impulsivity in adolescents with normal and excess weight. PloS One. 2012;7(11):e49185. doi:10.1371/journal.pone.0049185

75. Adise S, Ottino-Gonzalez J, Hayati Rezvan P, et al. Smaller subcortical volume relates to greater weight gain in girls with initially healthy weight. Obes Silver Spring Md. 2024;32(7):1389–1400. doi:10.1002/oby.24028

76. Zhou H, Hu Y, Li G, et al. Obesity is associated with progressive brain structural changes. Obesity. 2025;33(4):709–719. doi:10.1002/oby.24251

77. Manto M, Adamaszek M, Apps R, et al. Consensus Paper: Cerebellum and Reward. The Cerebellum. 2024;23(5):2169–2192. doi:10.1007/s12311-024-01702-0

78. Smith KS, Berridge KC. Opioid Limbic Circuit for Reward: Interaction between Hedonic Hotspots of Nucleus Accumbens and Ventral Pallidum. J Neurosci. 2007;27(7):1594–1605. doi:10.1523/JNEUROSCI.4205-06.2007

79. Adise S, Li ZA, Ottino-González J, Morys F, Chiarelli PA, Hershey T. Distinct Patterns of Weight Gain, Age, and Subcortical Microstructure in Early Adolescence. JAMA Netw Open. 2025;8(7):e2522211. doi:10.1001/jamanetworkopen.2025.22211

80. Li ZA, Samara A, Ray MK, et al. Childhood obesity is linked to putative neuroinflammation in brain white matter, hypothalamus, and striatum. Cereb Cortex Commun. 2023;4(2):tgad007. doi:10.1093/texcom/tgad007

81. Kaltenhauser S, Weber CF, Lin H, et al. Association of Body Mass Index and Waist Circumference With Imaging Metrics of Brain Integrity and Functional Connectivity in Children Aged 9 to 10 Years in the US, 2016-2018. JAMA Netw Open. 2023;6(5):e2314193. doi:10.1001/jamanetworkopen.2023.14193

82. Borowitz MA, Yokum S, Duval ER, Gearhardt AN. Weight-Related Differences in Salience, Default Mode, and Executive Function Network Connectivity in Adolescents. Obes Silver Spring Md. 2020;28(8):1438–1446. doi:10.1002/oby.22853

83. Geha P, Cecchi G, Todd Constable R, Abdallah C, Small DM. Reorganization of brain connectivity in obesity. Hum Brain Mapp. 2017;38(3):1403–1420. doi:10.1002/hbm.23462

84. Kelley WM, Wagner DD, Heatherton TF. In Search of a Human Self-Regulation System. Annu Rev Neurosci. 2015;38:389–411. doi:10.1146/annurev-neuro-071013-014243

85. Medic N, Kochunov P, Ziauddeen H, et al. BMI-related cortical morphometry changes are associated with altered white matter structure. Int J Obes 2005. 2019;43(3):523–532. doi:10.1038/s41366-018-0269-9

86. Verstynen TD, Weinstein AM, Schneider WW, Jakicic JM, Rofey DL, Erickson KI. Increased body mass index is associated with a global and distributed decrease in white matter microstructural integrity. Psychosom Med. 2012;74(7):682–690. doi:10.1097/PSY.0b013e318261909c

87. Kullmann S, Schweizer F, Veit R, Fritsche A, Preissl H. Compromised white matter integrity in obesity. Obes Rev Off J Int Assoc Study Obes. 2015;16(4):273–281. doi:10.1111/obr.12248

88. Xu J, Li Y, Lin H, Sinha R, Potenza MN. Body mass index correlates negatively with white matter integrity in the fornix and corpus callosum: a diffusion tensor imaging study. Hum Brain Mapp. 2013;34(5):1044–1052. doi:10.1002/hbm.21491

89. Alarcón G, Ray S, Nagel BJ. Lower Working Memory Performance in Overweight and Obese Adolescents Is Mediated by White Matter Microstructure. J Int Neuropsychol Soc JINS. 2016;22(3):281–292. doi:10.1017/S1355617715001265

90. Yau PL, Kang EH, Javier DC, Convit A. Preliminary evidence of cognitive and brain abnormalities in uncomplicated adolescent obesity. Obes Silver Spring Md. 2014;22(8):1865–1871. doi:10.1002/oby.20801

91. Ou X, Andres A, Pivik RT, Cleves MA, Badger TM. Brain gray and white matter differences in healthy normal weight and obese children. J Magn Reson Imaging JMRI. 2015;42(5):1205–1213. doi:10.1002/jmri.24912

92. Albertsson-Wikland K, Niklasson A, Gelander L, Holmgren A, Nierop AFM. Novel type of references for BMI aligned for onset of puberty - using the QEPS growth model. BMC Pediatr. 2022;22(1):238. doi:10.1186/s12887-022-03304-3

93. Huang A, Reinehr T, Roth CL. Connections Between Obesity and Puberty: Invited by Manuel Tena-Sempere, Cordoba. Curr Opin Endocr Metab Res. 2020;14:160–168. doi:10.1016/j.coemr.2020.08.004

94. Morys F, Tremblay C, Rahayel S, et al. Neural correlates of obesity across the lifespan. Commun Biol. 2024;7(1):656. doi:10.1038/s42003-024-06361-9

95. Fernández-Andújar M, Morales-García E, García-Casares N. Obesity and Gray Matter Volume Assessed by Neuroimaging: A Systematic Review. Brain Sci. 2021;11(8):999. doi:10.3390/brainsci11080999

96. Gómez-Apo E, Mondragón-Maya A, Ferrari-Díaz M, Silva-Pereyra J. Structural Brain Changes Associated with Overweight and Obesity. J Obes. 2021;2021:6613385. doi:10.1155/2021/6613385

97. Sharkey RJ, Karama S, Dagher A. Overweight is not associated with cortical thickness alterations in children. Front Neurosci. 2015;9:24. doi:10.3389/fnins.2015.00024

98. Johnson WE, Li C, Rabinovic A. Adjusting batch effects in microarray expression data using empirical Bayes methods. Biostat Oxf Engl. 2007;8(1):118–127. doi:10.1093/biostatistics/kxj037

99. Zhu X, Kim Y, Ravid O, et al. Neuroimaging-based classification of PTSD using data-driven computational approaches: A multisite big data study from the ENIGMA-PGC PTSD consortium. NeuroImage. 2023;283:120412. doi:10.1016/j.neuroimage.2023.120412

100. Kim BG, Kim G, Abe Y, et al. White matter diffusion estimates in obsessive-compulsive disorder across 1653 individuals: machine learning findings from the ENIGMA OCD Working Group. Mol Psychiatry. 2024;29(4):1063–1074. doi:10.1038/s41380-023-02392-6

