## Supplemental figures S1-S13 and Supplemental tables S1-S2 for "Brain, genetic and demographic factors predict current body fat estimate and weight gain in (pre)adolescents: evidence from the ABCD study"

### Supplemental results

**Figure S1.** Mean ElasticNet regression model coefficients for sMRI brain imaging measures not visualizable by brainplot.

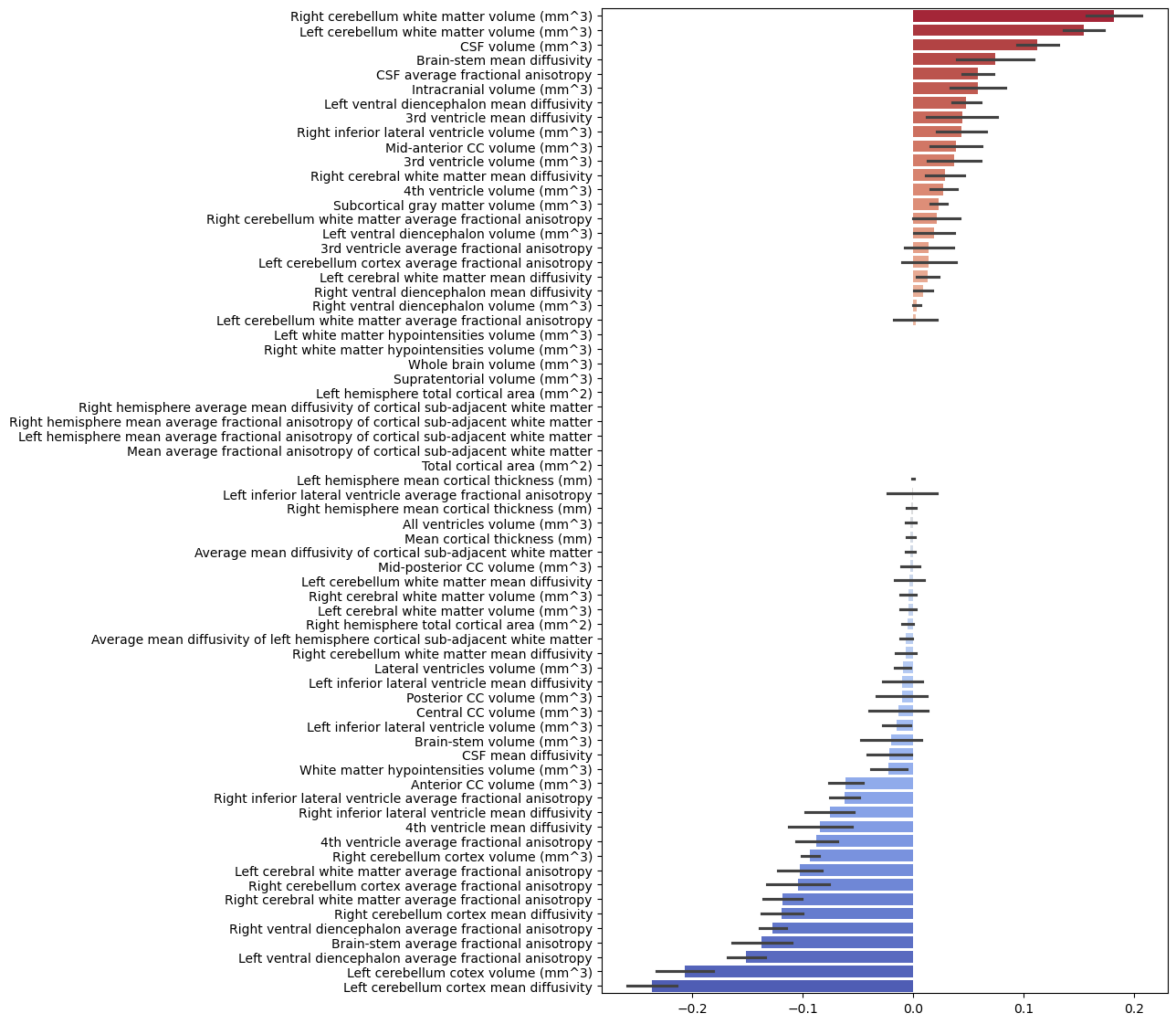

**Figure S2.** Mean ElasticNet regression model coefficients for sMRI non-imaging features.

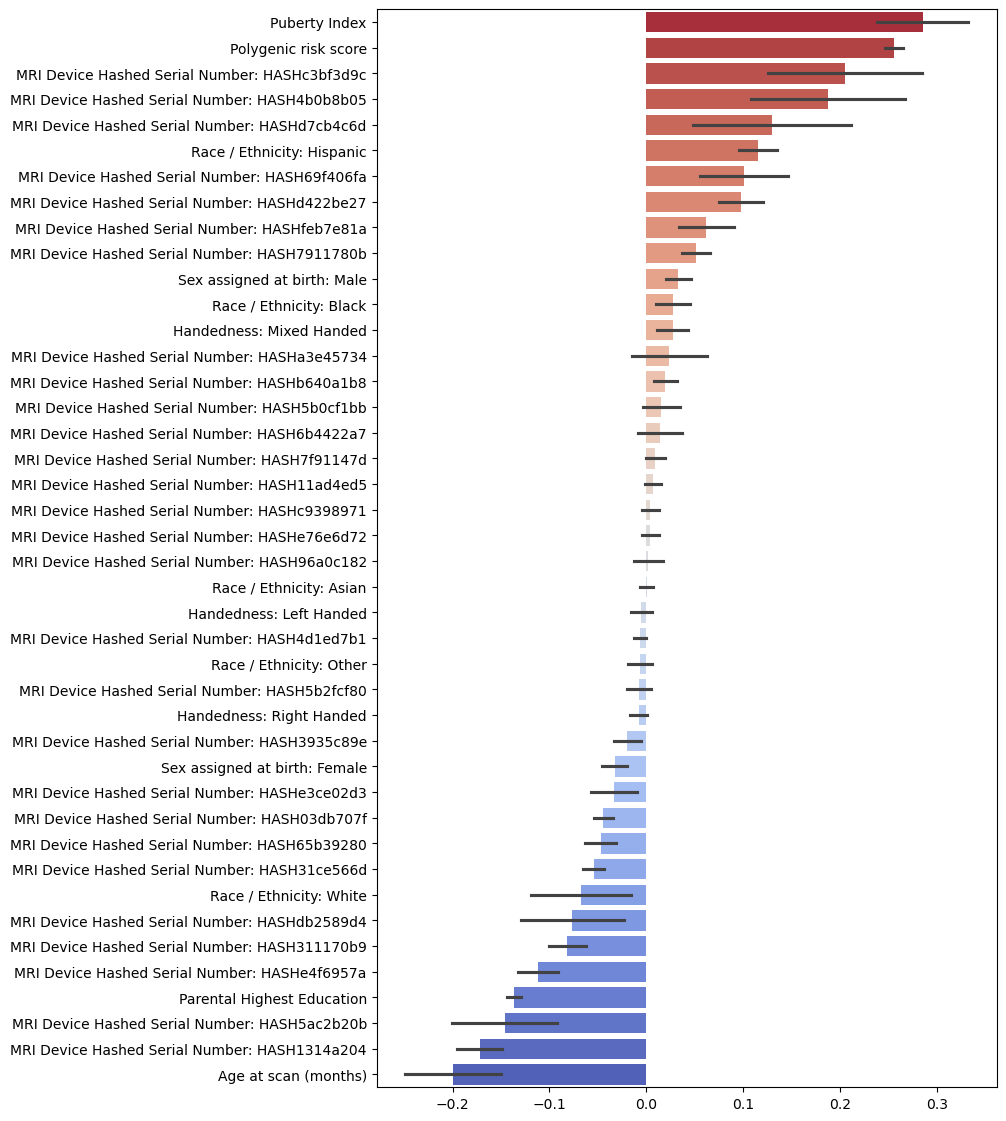

**Figure S3**. A piechart showing the relative contributions of different categories of features in terms of their absolute aggregate coefficients for sMRI regression model
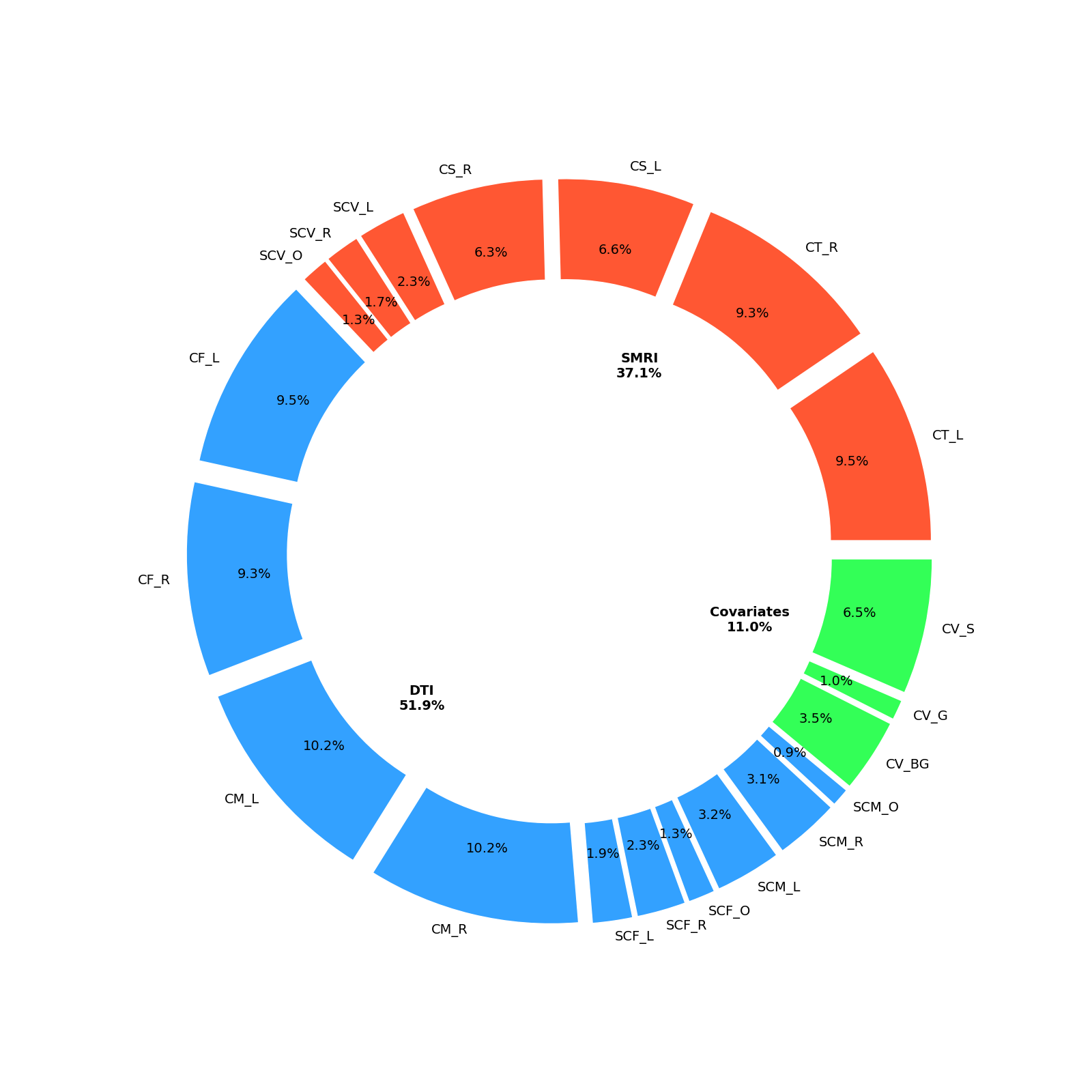

**Figure S4.** Mean ElasticNet regression model coefficients for rs-fMRI non-imaging features.

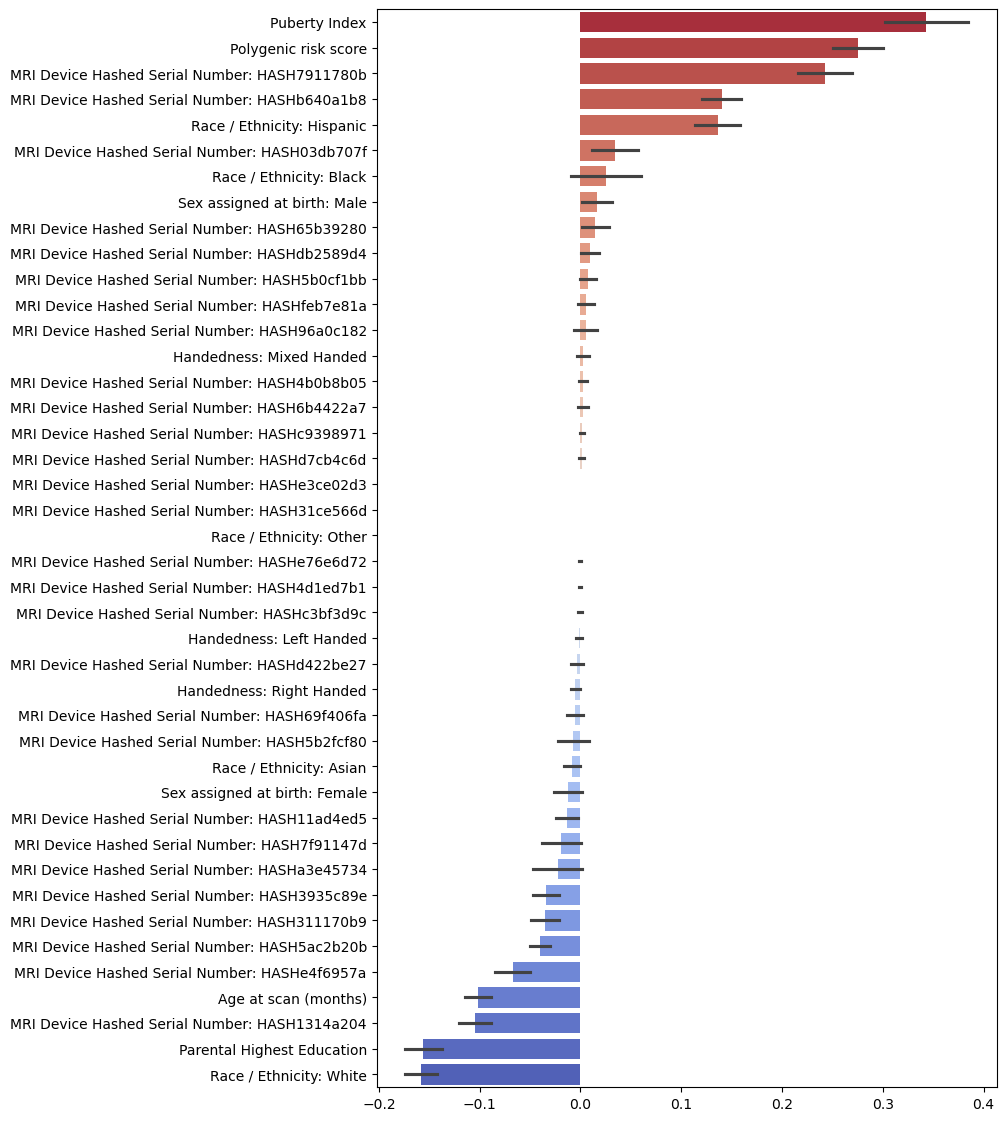

**Figure S5**. A piechart showing the relative contributions of different categories of features in terms of their absolute aggregate coefficients for fMRI regression model.

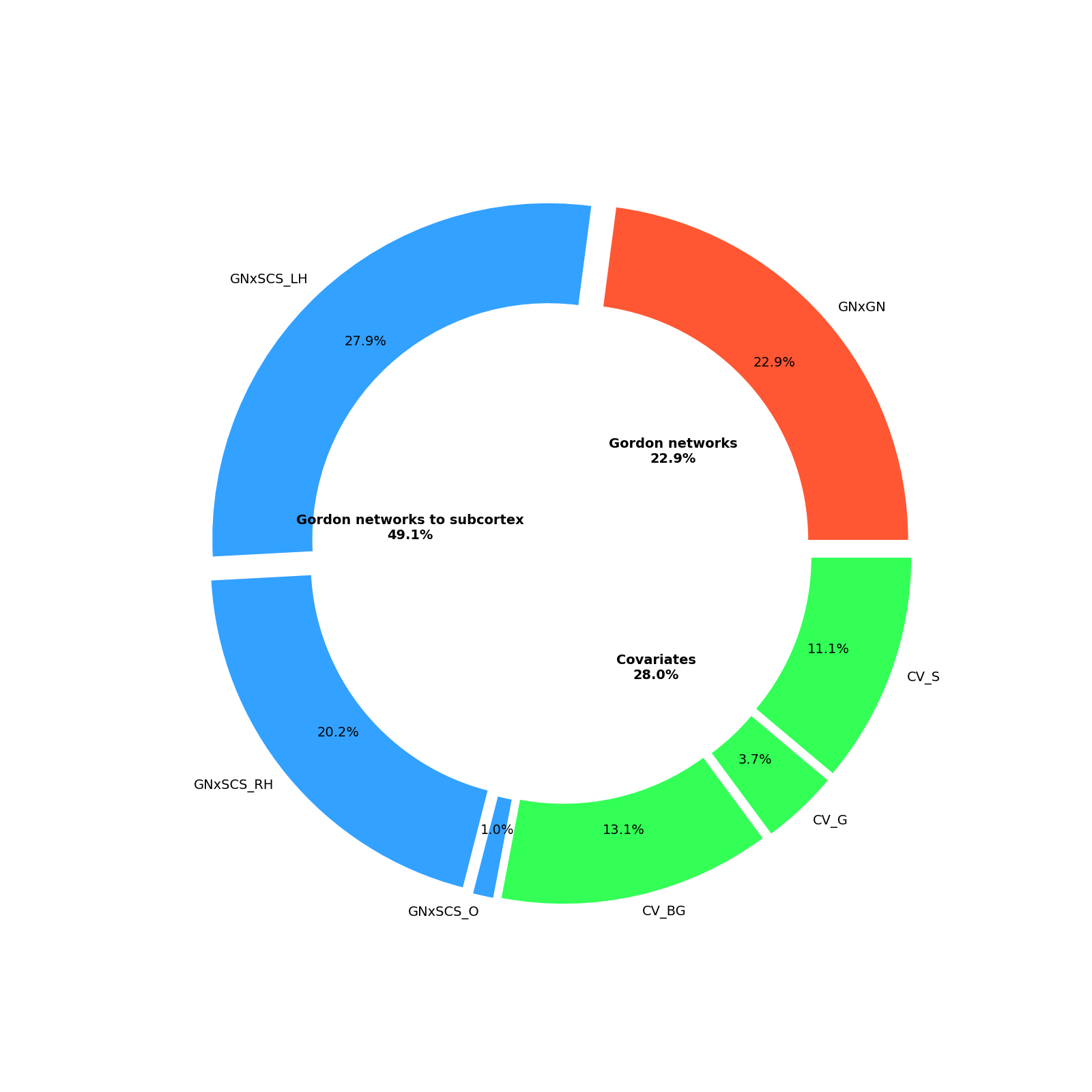

**Figure S6.** Mean ElasticNet classification model coefficients for sMRI brain imaging measures not visualizable by brainplot.

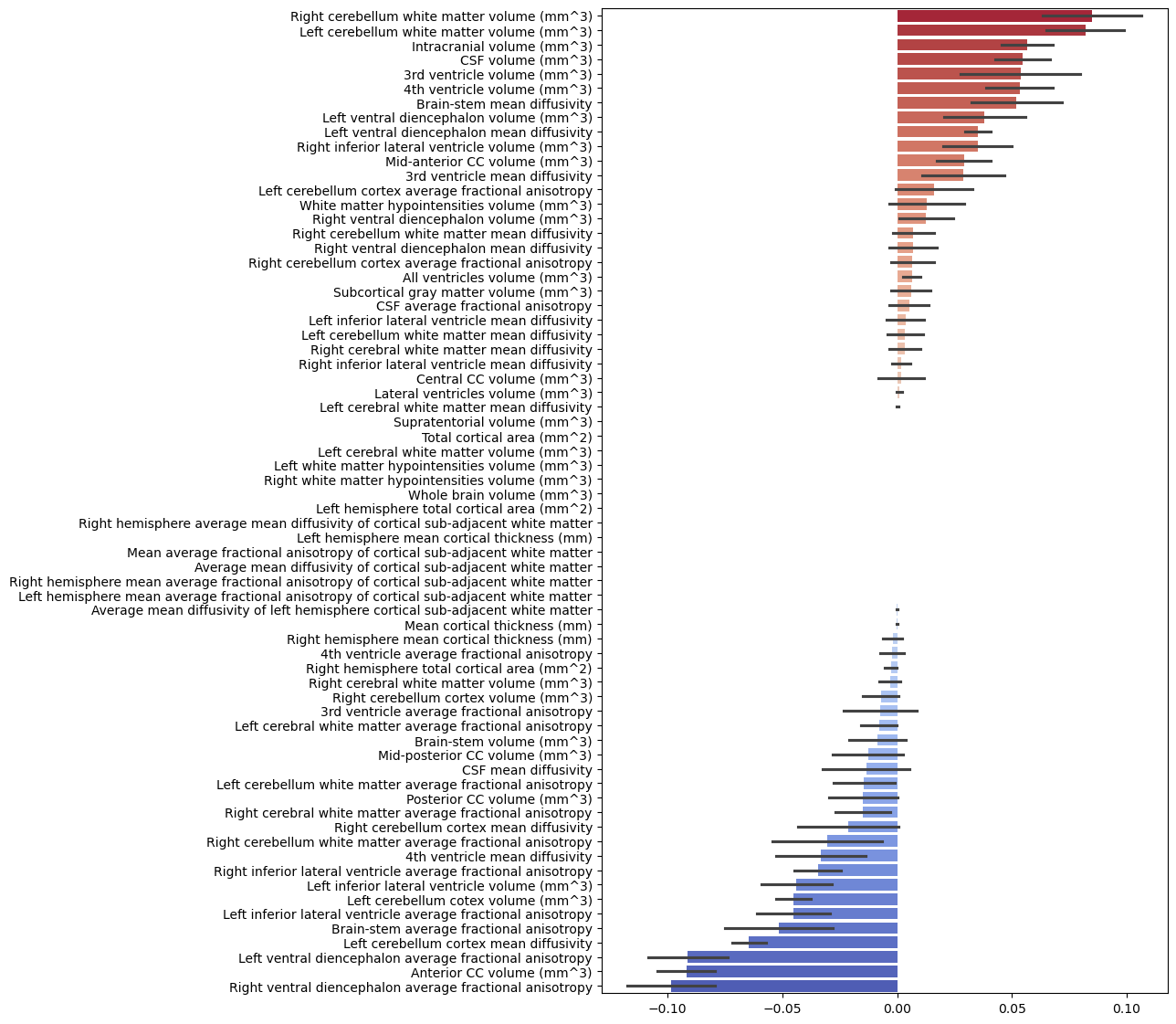

**Figure S7.** Mean ElasticNet classification model coefficients for sMRI non-imaging features.

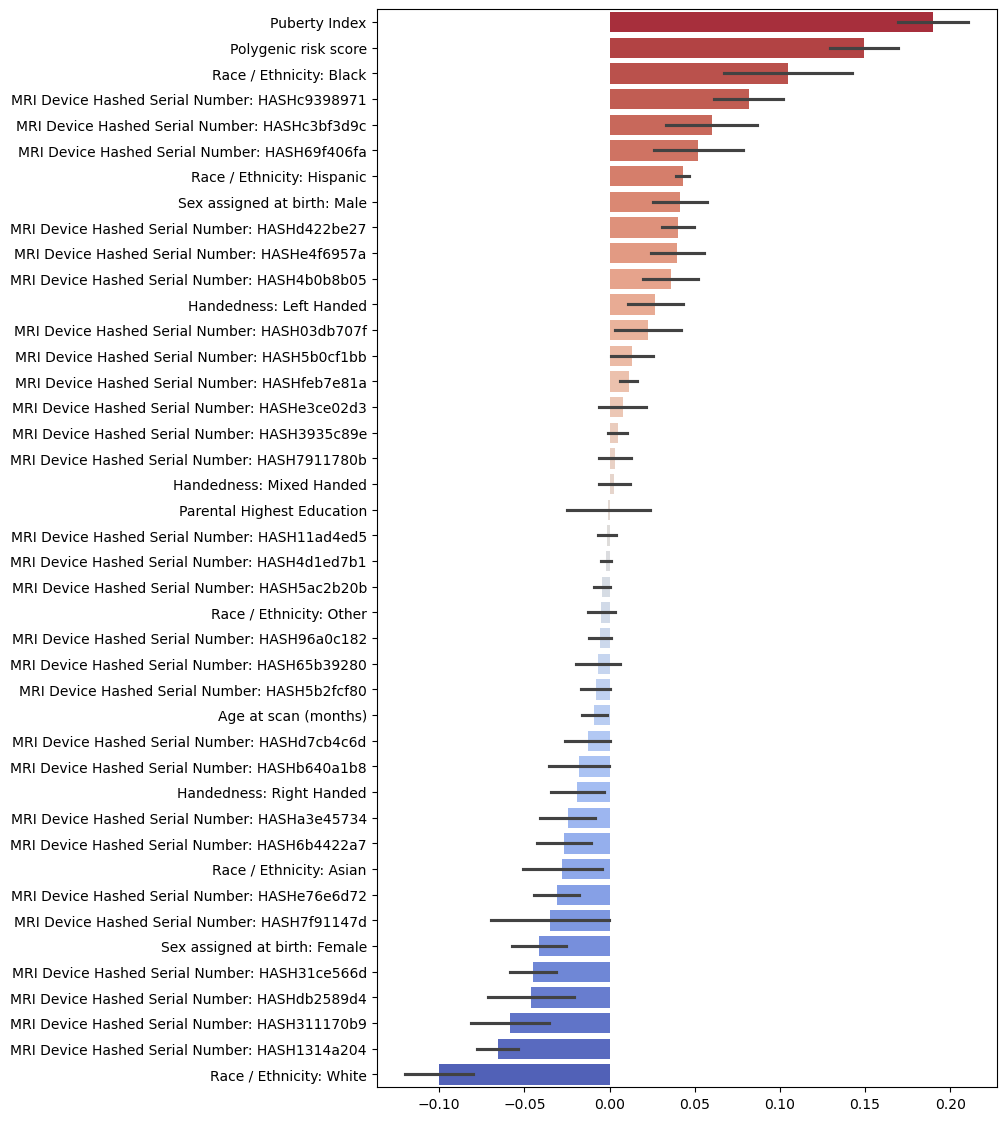

**Figure S8**. A piechart showing the relative contributions of different categories of features in terms of their absolute aggregate coefficients for sMRI classification model.

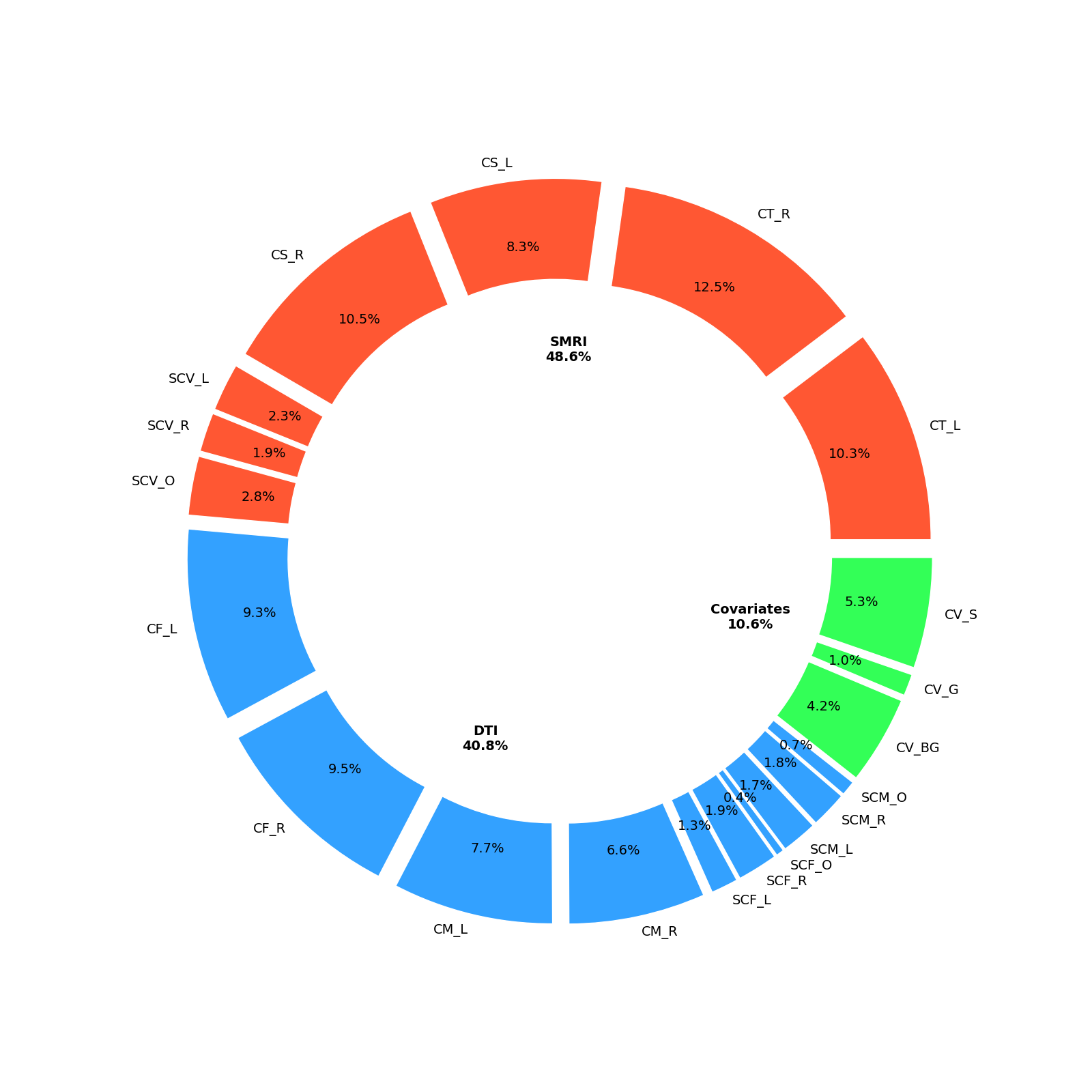

**Figure S9.** Mean ElasticNet classification model coefficients for rs-fMRI non-imaging features.

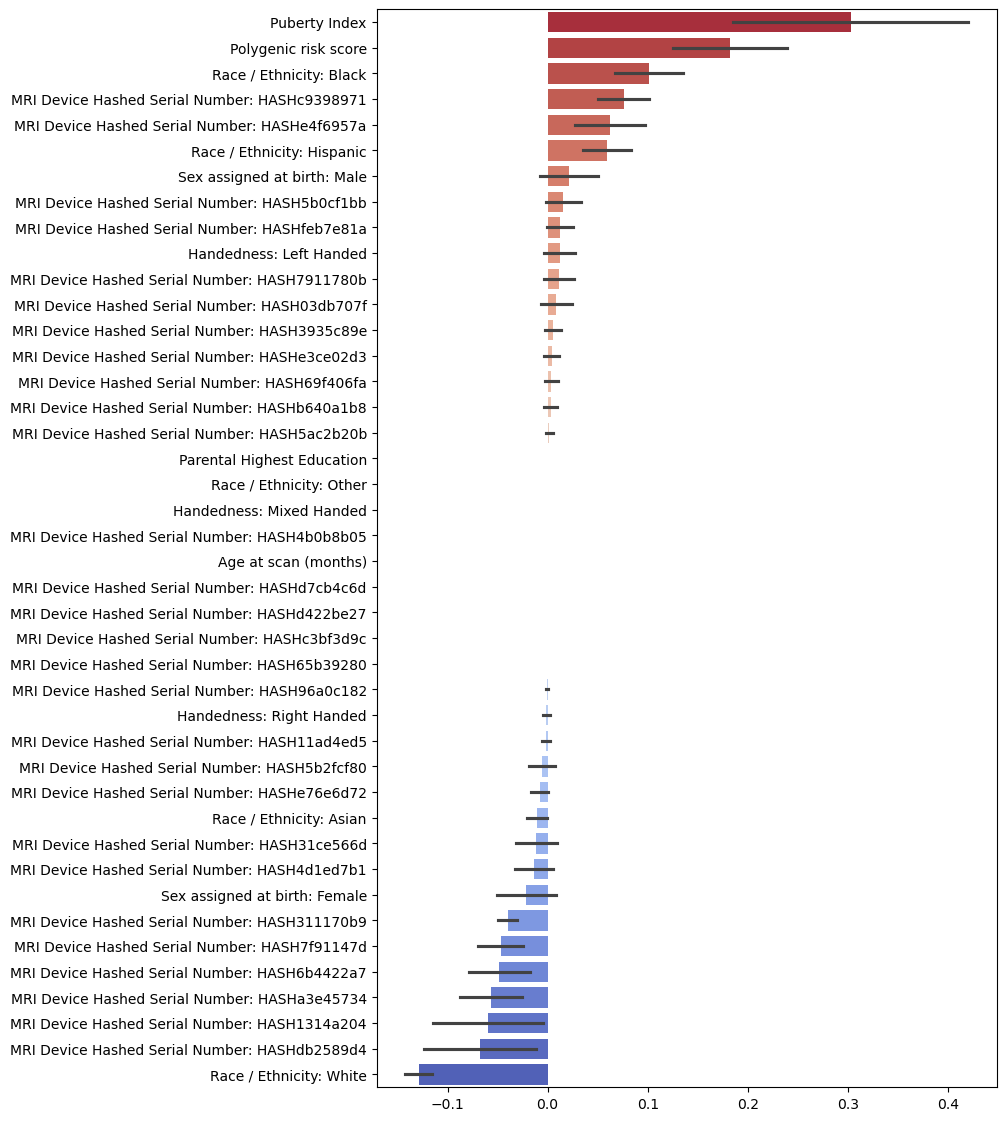

**Figure S10**. A piechart showing the relative contributions of different categories of features in terms of their absolute aggregate coefficients for fMRI classification model.

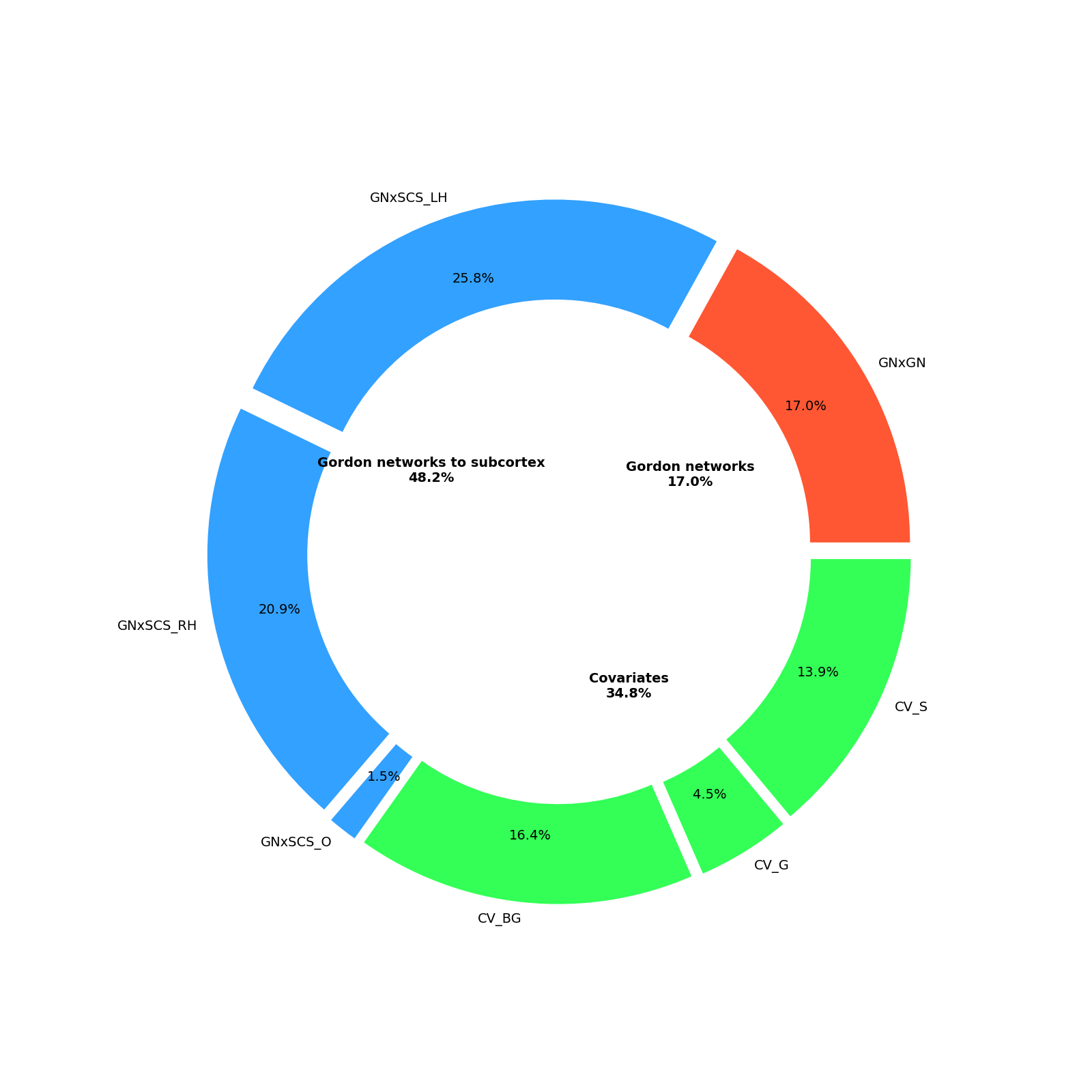

**Figure S11.** Principal components for cortical thickness, surface area, and subcortical volume

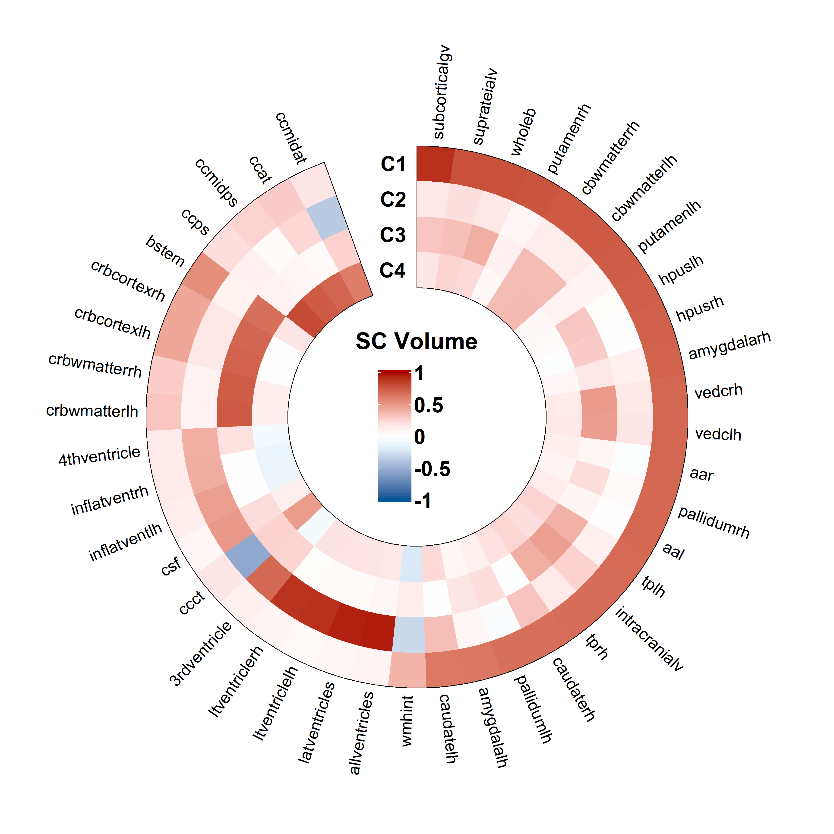

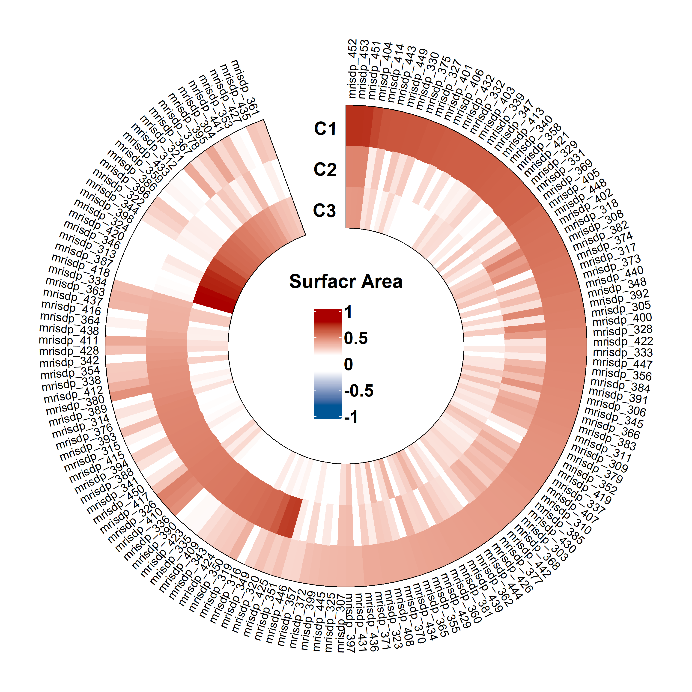

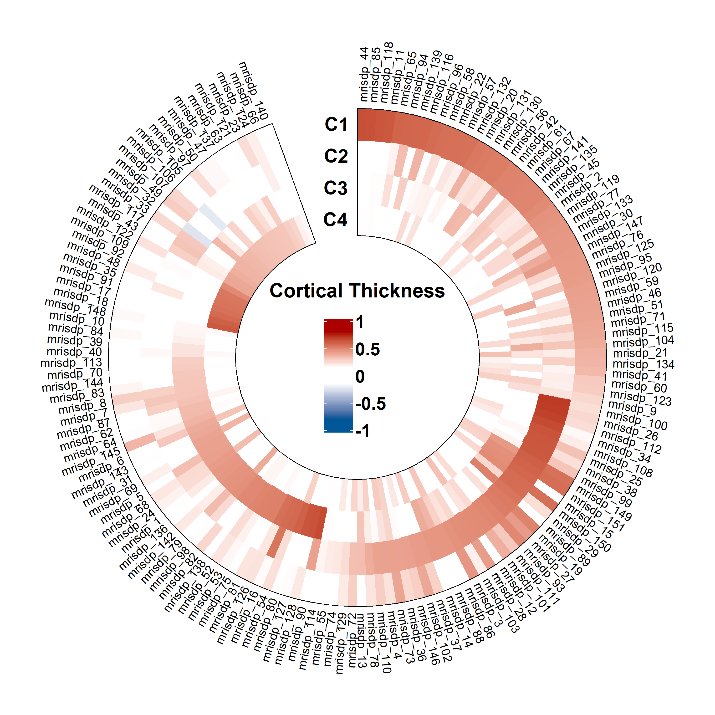

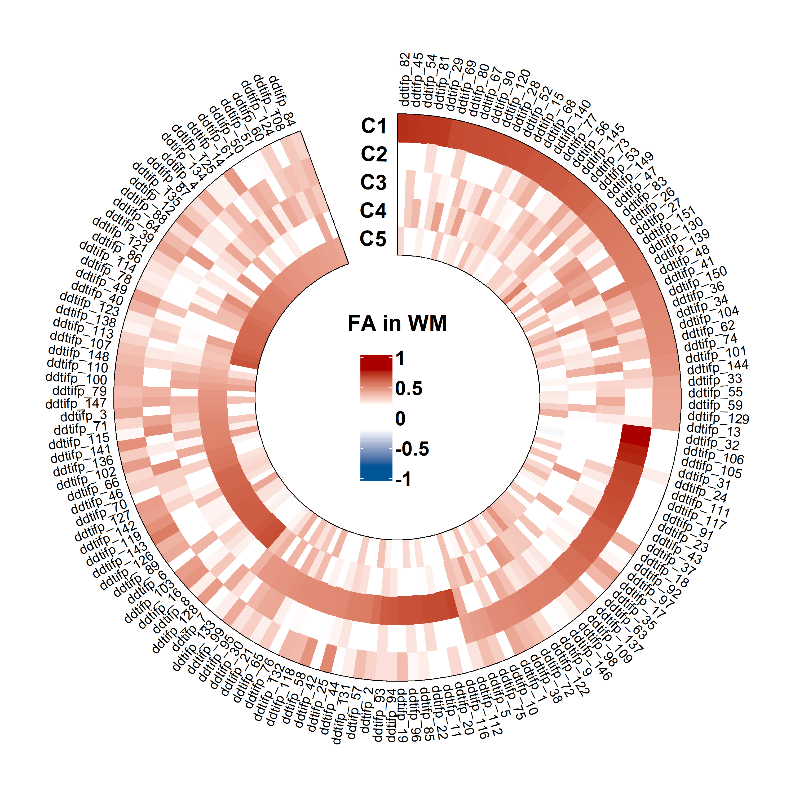

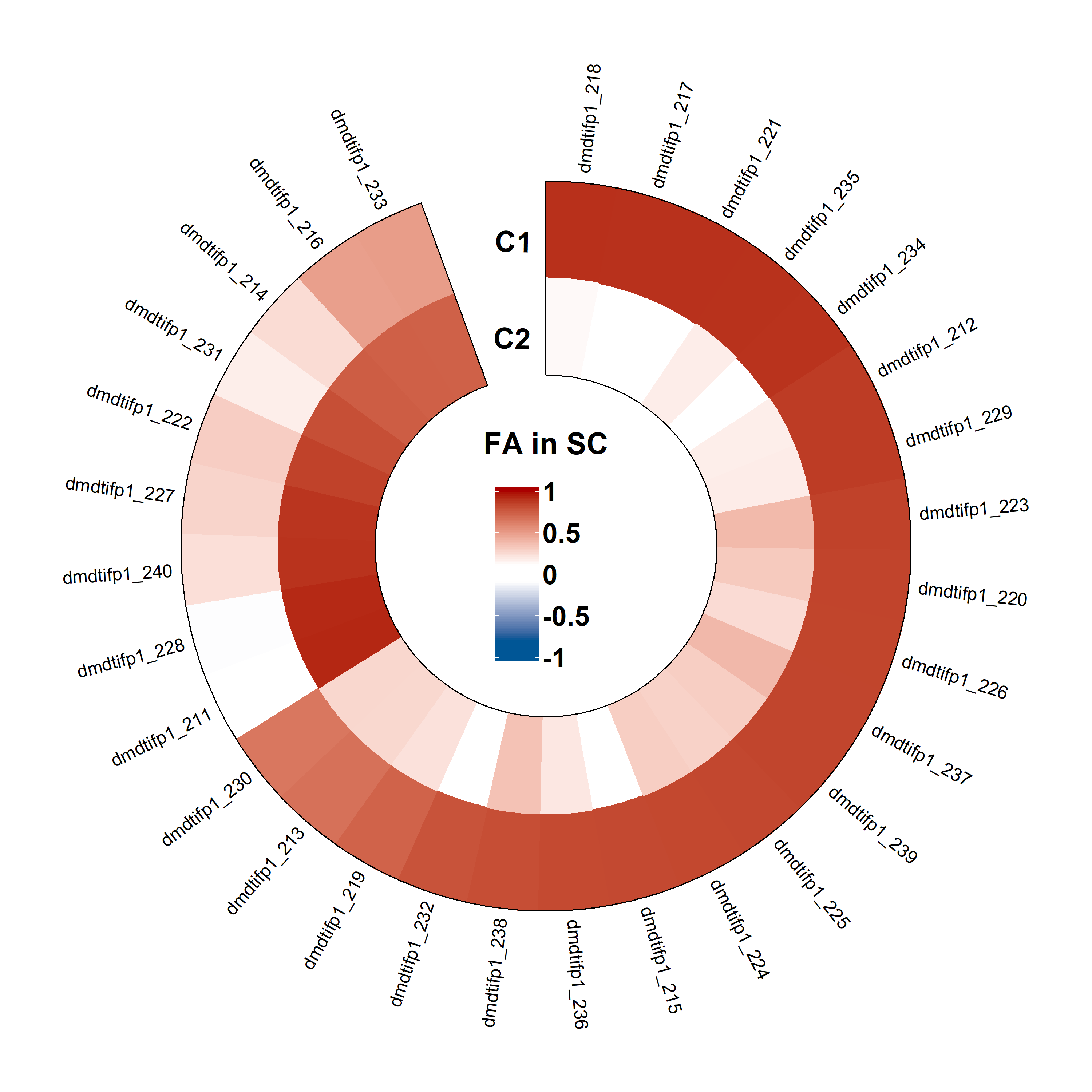

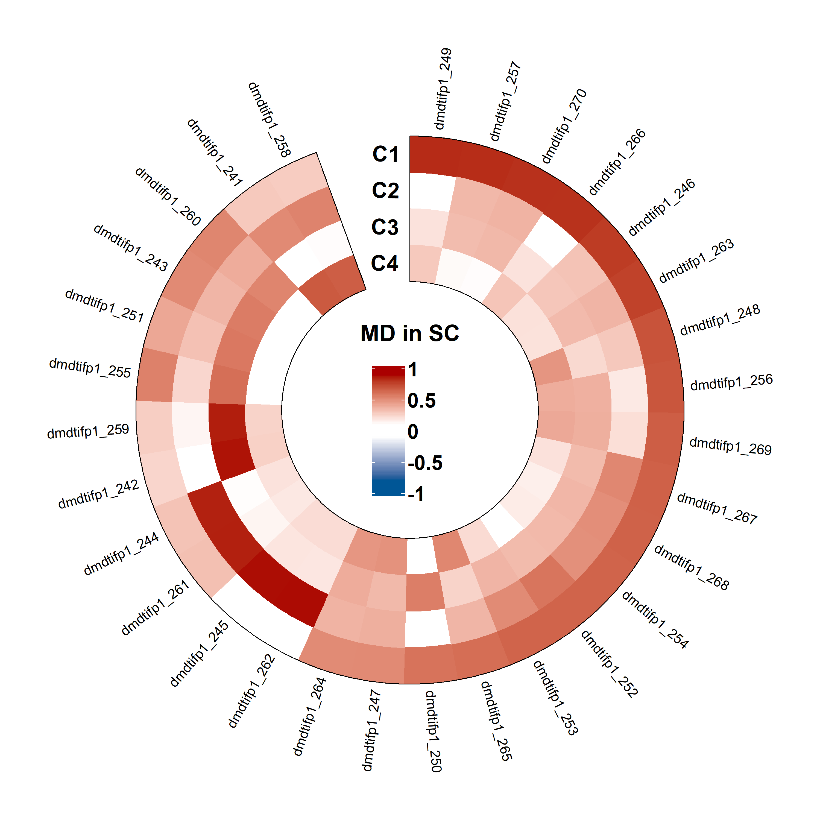
**Figure S12.** Principal components for FA/MD in white matter and in subcortical regions

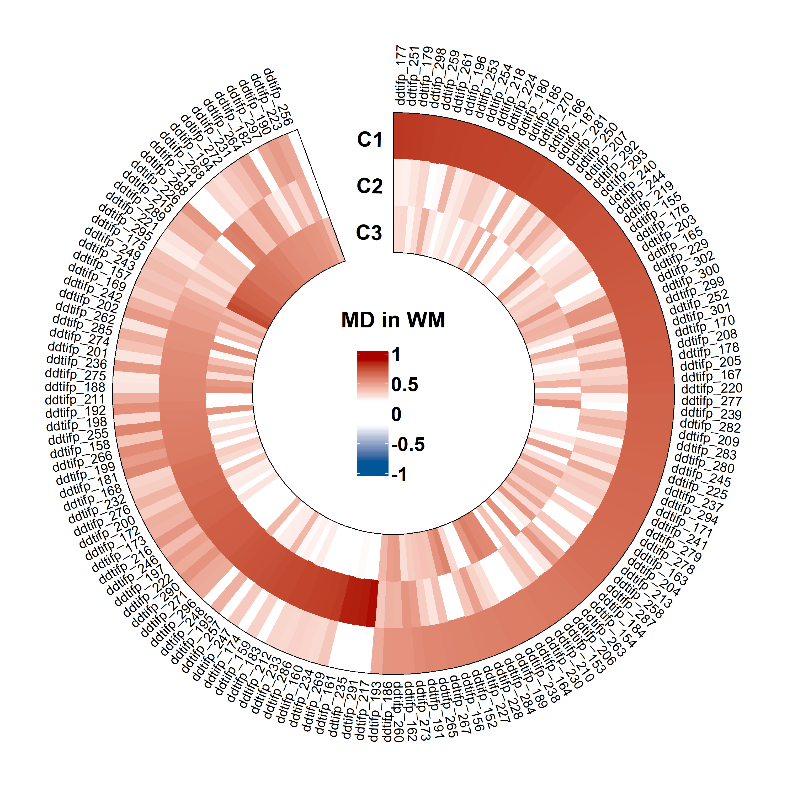

**Figure S13.** Principal components for rsfMRI (subcortical inter- & intra-network, and cortical-to-subcortical connectivity)

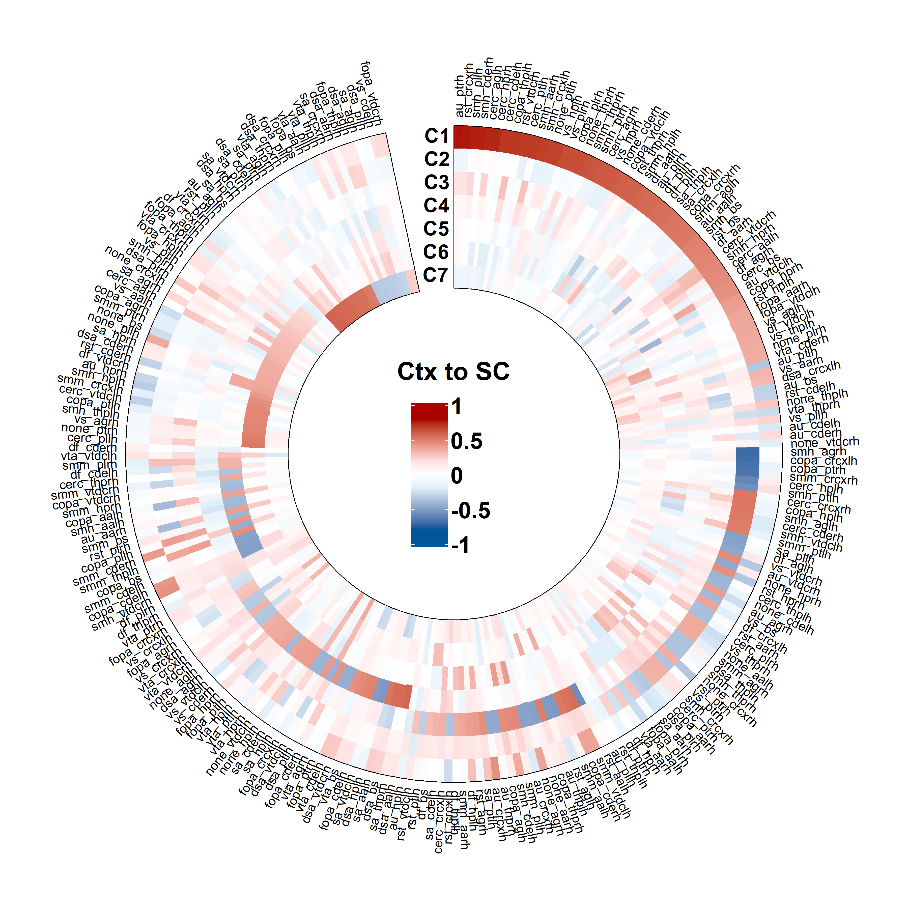

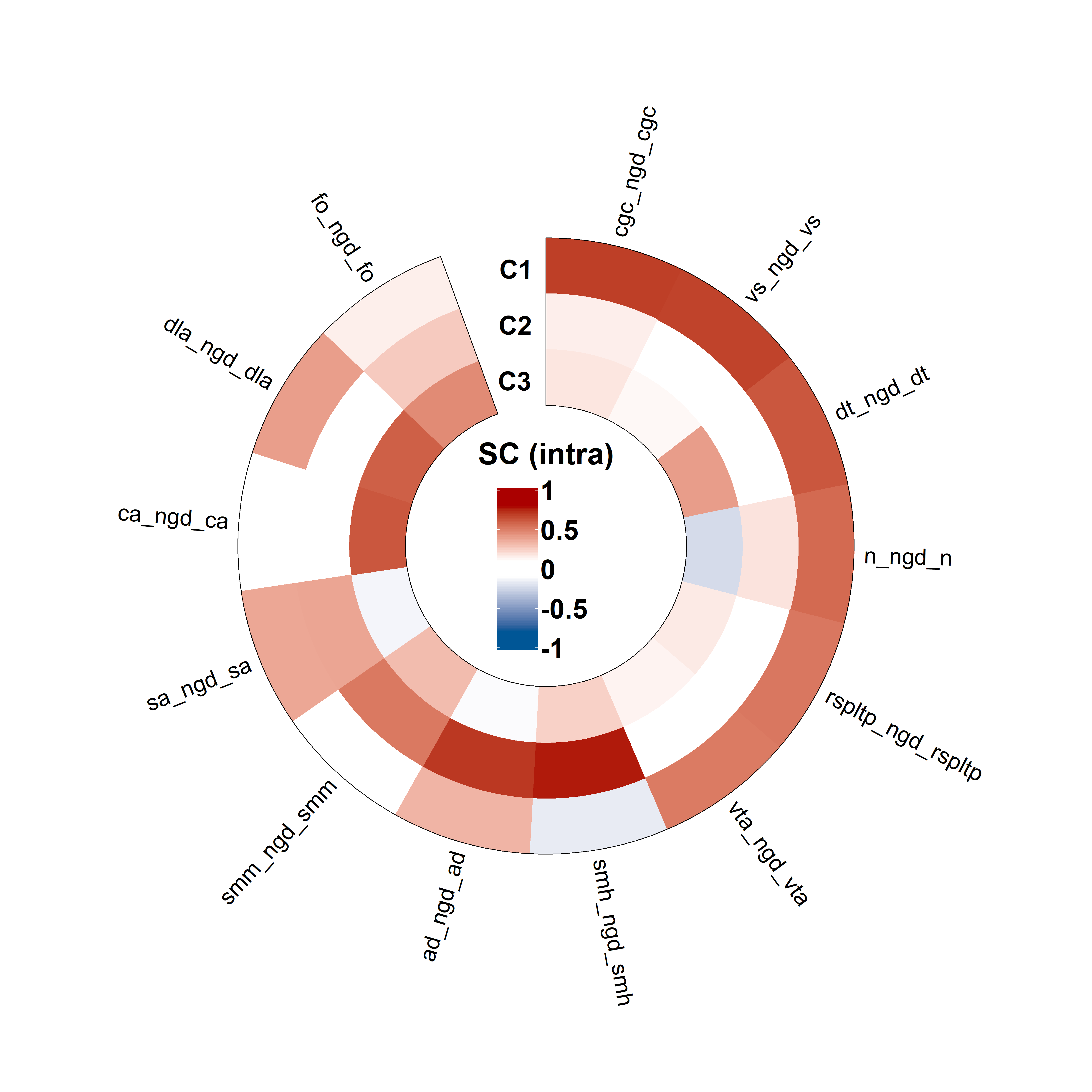

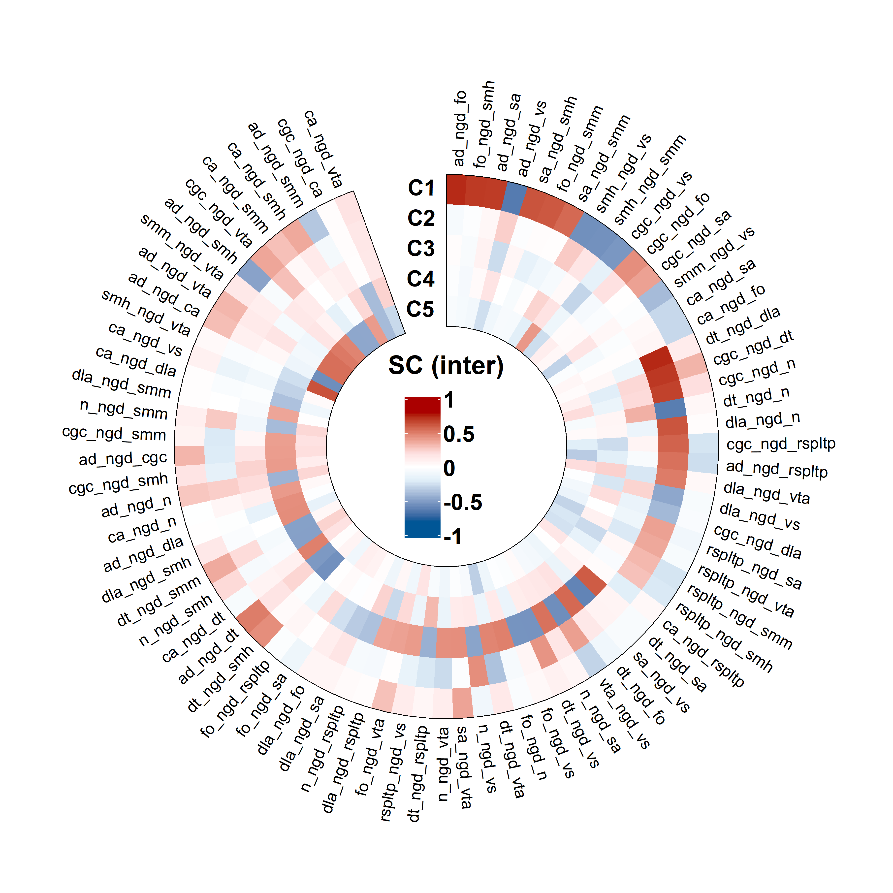

**Table S1:** Results of the leave-one-site-out cross-validation analysis for brain imaging measures only datasets. Test site sample size (N) and cross-validated performance metric reported for each test site separately in terms of R^2^ for the regression task and ROC-AUC for the classification task and averaged over the sites. Weighted mean score computed by weighing the test-site specific scores with site sample size.

| **Test site ID** | **sMRI reg** | | **fMRI reg** | | **sMRI clf** | | **fMRI clf** | |
| --- | --- | --- | --- | --- | --- | --- | --- | --- |
|  | N | R^2^ | N | R^2^ | N | AUC | N | AUC |
| G010 | 179 | -0.217 | 170 | 0.081 | 78 | 0.786 | 73 | 0.533 |
| G031 | 429 | 0.192 | 407 | 0.105 | 157 | 0.698 | 150 | 0.525 |
| G032 | 424 | 0.128 | 443 | 0.076 | 161 | 0.743 | 171 | 0.658 |
| G075 | 425 | -0.129 | 428 | 0.102 | 139 | 0.672 | 146 | 0.671 |
| G087 | 228 | 0.054 | 228 | -0.016 | 78 | 0.598 | 75 | 0.568 |
| P023 | 226 | 0.143 | 205 | -0.088 | 104 | 0.732 | 91 | 0.584 |
| P043 | 394 | -0.003 | 318 | -0.020 | 146 | 0.614 | 118 | 0.594 |
| P064 | 214 | 0.191 | 184 | 0.054 | 58 | 0.842 | 50 | 0.573 |
| S011 | 404 | 0.185 | 387 | -0.018 | 126 | 0.760 | 118 | 0.569 |
| S012 | 396 | 0.245 | 345 | 0.046 | 124 | 0.737 | 103 | 0.448 |
| S013 | 256 | 0.133 | 225 | -0.103 | 70 | 0.667 | 61 | 0.691 |
| S014 | 727 | 0.205 | 716 | -0.003 | 242 | 0.760 | 236 | 0.604 |
| S020 | 308 | 0.147 | 274 | -0.044 | 148 | 0.692 | 134 | 0.568 |
| S021 | 337 | 0.215 | 304 | 0.043 | 147 | 0.749 | 132 | 0.642 |
| S022 | 285 | -0.162 | 275 | -0.208 | 119 | 0.773 | 112 | 0.495 |
| S042 | 399 | 0.281 | 361 | 0.044 | 155 | 0.770 | 145 | 0.617 |
| S053 | 374 | 0.098 | 343 | -0.020 | 125 | 0.680 | 117 | 0.676 |
| S065 | 285 | 0.215 | 256 | 0.122 | 94 | 0.780 | 85 | 0.695 |
| S076 | 306 | 0.280 | 276 | 0.124 | 109 | 0.641 | 96 | 0.646 |
| S086 | 260 | 0.140 | 231 | 0.007 | 94 | 0.751 | 82 | 0.515 |
| S090 | 227 | 0.335 | 193 | 0.134 | 77 | 0.804 | 63 | 0.666 |
| Mean (SD) | 7083 | 0.127 (0.143) | 6569 | 0.020 (0.084) | 2551 | 0.726 (0.062) | 2358 | 0.597 (0.067) |
| Weighted mean |  | 0.136 |  | 0.022 |  | 0.724 |  | 0.598 |

**Table S2:** Results of the leave-one-site-out cross-validation analysis for brain imaging and non-imaging measures datasets. Test site sample size (N) and cross-validated performance metric reported for each test site separately in terms of R^2^ for the regression task and ROC-AUC for the classification task and averaged over the sites. Weighted mean score computed by weighing the test-site specific scores with site sample size.

| **Test site ID** | **sMRI reg** | | **fMRI reg** | | **sMRI clf** | | **fMRI clf** | |
| --- | --- | --- | --- | --- | --- | --- | --- | --- |
|  | N | R^2^ | N | R^2^ | N | AUC | N | AUC |
| G010 | 179 | -0.149 | 170 | 0.087 | 78 | 0.825 | 73 | 0.702 |
| G031 | 429 | 0.250 | 407 | 0.224 | 157 | 0.721 | 150 | 0.690 |
| G032 | 424 | 0.192 | 443 | 0.137 | 161 | 0.755 | 171 | 0.688 |
| G075 | 425 | -0.050 | 428 | 0.184 | 139 | 0.702 | 146 | 0.710 |
| G087 | 228 | 0.127 | 228 | 0.113 | 78 | 0.645 | 75 | 0.747 |
| P023 | 226 | 0.217 | 205 | 0.013 | 104 | 0.752 | 91 | 0.663 |
| P043 | 394 | 0.051 | 318 | 0.034 | 146 | 0.624 | 118 | 0.610 |
| P064 | 214 | 0.235 | 184 | 0.116 | 58 | 0.829 | 50 | 0.550 |
| S011 | 404 | 0.219 | 387 | 0.035 | 126 | 0.768 | 118 | 0.616 |
| S012 | 396 | 0.247 | 345 | 0.054 | 124 | 0.715 | 103 | 0.535 |
| S013 | 256 | 0.133 | 225 | -0.070 | 70 | 0.653 | 61 | 0.674 |
| S014 | 727 | 0.263 | 716 | 0.126 | 242 | 0.795 | 236 | 0.731 |
| S020 | 308 | 0.173 | 274 | 0.058 | 148 | 0.709 | 134 | 0.646 |
| S021 | 337 | 0.236 | 304 | 0.146 | 147 | 0.752 | 132 | 0.684 |
| S022 | 285 | -0.111 | 275 | -0.155 | 119 | 0.794 | 112 | 0.651 |
| S042 | 399 | 0.329 | 361 | 0.124 | 155 | 0.783 | 145 | 0.720 |
| S053 | 374 | 0.134 | 343 | 0.064 | 125 | 0.679 | 117 | 0.719 |
| S065 | 285 | 0.237 | 256 | 0.133 | 94 | 0.768 | 85 | 0.749 |
| S076 | 306 | 0.299 | 276 | 0.182 | 109 | 0.642 | 96 | 0.610 |
| S086 | 260 | 0.171 | 231 | 0.008 | 94 | 0.783 | 82 | 0.670 |
| S090 | 227 | 0.376 | 193 | 0.211 | 77 | 0.835 | 63 | 0.768 |
| Mean (SD) |  | 0.170 (0.133) |  | 0.087 (0.089) |  | 0.739 (0.063) |  | 0.673 (0.061) |
| Weighted mean |  | 0.180 |  | 0.095 |  | 0.739 |  | 0.677 |
